# Genetic Risk for High Body Mass Index Before and Amidst the Obesity Epidemic: Cross-Cohort Analysis of Four British Birth Cohort Studies

**DOI:** 10.1101/2024.10.24.24315860

**Authors:** Liam Wright, Neil M Davies, Gemma Shireby, Dylan M Williams, Tim T Morris, David Bann

## Abstract

Obesity is a highly heritable trait, but rising obesity rates over the past five decades suggest environmental change is also of profound importance. We conducted a cross-cohort analysis to examine how associations between genetic risk for high BMI and observed BMI differed in four British birth cohorts born before and amidst the obesity epidemic (1946, 1958, 1970 and ∼2001, respectively; N = 19,379). BMI (kg/m^2^) was measured at multiple time points between ages 3 and 69 years. We used polygenic indices (PGI) derived from GWAS of adulthood and childhood BMI, respectively, with mixed effects models used to estimate associations with mean BMI and quantile regression used to assess associations across the distribution of BMI. We further used Genomic Relatedness Restricted Maximum Likelihood (GREML) to calculate SNP-heritability (SNP-h^2^) at each age. Adulthood BMI PGI was associated with BMI in all cohorts and ages but was more strongly associated with BMI in more recently born generations. For example, at age 16y, a 1 SD increase in the adulthood PGI was associated with 0.43 kg/m^2^ (0.34, 0.51) higher BMI in the 1946c and 0.90 kg/m^2^ (0.83, 0.97) higher BMI in the 2001c. Cross-cohort differences widened with age and were larger at the upper end of the BMI distribution, indicating disproportionate increases in obesity in more recent generations for those with higher PGIs. Differences were also observed when using the childhood PGI, but there were no clear, consistent differences in SNP-h^2^. Findings highlight how the environment can modify genetic influence; genetic effects on BMI differed by birth cohort, age, and outcome centile.

## Introduction

Obesity is a leading cause of morbidity and premature mortality worldwide [1], with the global economic cost of overweight and obesity estimated to exceed $2tn per annum [2]. More than one in four adults and one in five 11-year-olds in England is obese [3]. The strong tracking of body mass index (BMI) across the life course [4] raises the possibility that current generations will spend more time obese [5], increasing the risk of public health problems in years to come [6].

It was not always thus. The prevalence of obesity has increased dramatically in industrialised nations over the past five decades, though the timing and extent of this increase has differed markedly across countries [7]. In England, obesity rates among children and adults have more than tripled since the mid-1970s [8–11]. The precipitous increase in obesity, faster than any plausible genetic change at a population level, suggests an important role of the environment in determining body weight.

Obesity is likely proximally caused by an imbalance between energy consumed and energy expended. Multiple societal changes have occurred alongside the obesity epidemic that are thought to have negatively influenced energy balance, though the relative contribution of changes in energy consumption and expenditure remains debated [10, 12–15]. Technological, economic and social developments have progressively ‘engineered’ physical effort out of many people’s lives [16], for instance, through the introduction of labour-saving devices in the home and in the workplace, the growth of sedentary leisure activities, such as television and video games, and the decline in manual employment [17]. Food, especially sugary and fatty food, has also become cheaper [16], the relative share of food expenditure on processed foodstuffs has increased [18], and fast-food outlets have expanded in number [19].

Yet, while obesity rates have increased, the underlying change in the distribution of body mass index (BMI) has not been uniform. Instead, the population distribution of BMI has become more variable and more skewed – levels of underweight are almost unchanged, while the increase in median BMI has been small relative to the growth in obesity rates [5, 8, 9, 20]. This change in the distribution of BMI suggests that individuals differ in their susceptibility to the obesogenic environment. One source of these differences may be genetics. BMI is highly heritable, with estimates from twin studies ranging 47-90% [21], and genetic variants that increase the risk of obesity can operate through the environment [22]. For instance, variants of the *FTO* gene strongly related to obesity risk [23] are also associated with multiple behavioural and psychological dispositions related to eating, such as increased hunger and lower satiety [24]. These dispositions may be more likely to translate into higher BMI in conditions where energy-dense food is cheap, salient, and widely available and where individuals are unlikely to compensate by increasing physical activity – hallmarks of the obesity epidemic [22, 25].

Several gene-environment interaction (GxE) studies have investigated whether heritability and genotypic penetrance – defined as the association between genotype and the level of a trait (in this case, BMI) – have increased alongside the obesity epidemic. These proxy for exposure to the obesity epidemic by birth year or year of assessment [26–34], prevalence of obesity or mean BMI in the sample studied [35, 36]. These studies show that, while average BMI has increased regardless of genotype [26], genetic penetrance has grown alongside the obesity epidemic, while heritability has stayed relatively stable; between-person differences in BMI according to genotype are now increased, but the *proportion* of variation in BMI explained is largely unchanged.

However, a limitation of these studies is that they have almost exclusively used data from adults – particularly older adults from the US – rather than children or adolescents. This is important as there are distinct genetic effects at different developmental stages [37]; two PGIs trained on adult and childhood BMI, respectively, are only correlated at r ∼ 0.4 [38]. Further, the environments people encounter that are relevant for obesity change as individuals develop in ways that could influence genetic effects. For instance, young children are generally given less agency over the food they consume and have very different physical activity levels than adults. It is therefore unclear to what extent previous results generalize to younger age groups. Further, most use regional rather than national samples.

Previous studies have also focused on changes in the association of genetics and mean BMI or obesity, specifically, rather than investigating changing associations across the full distribution of BMI. As noted, the obesity epidemic is marked by increasing skewness in BMI [8, 20]. Previous research has shown that an adult BMI PGI is particularly associated with high levels of BMI (e.g., Class II obesity) [38], but this has not been examined comparing cohorts differentially exposed to the obesity epidemic.

The British Birth Cohort Studies [39], which follow cohorts of individuals born in 1946, 1958, 1970, and 2000/02, offer a unique window into changing obesity rates. The cohorts have recently been genotyped and have multiple measurements of BMI across life, collectively spanning pre- and post-obesity epidemic periods. The oldest cohort grew up in a uniquely relatively uniform food environment and were eight years old when post-World War II food rationing ended in the UK [40, 41], while the youngest cohort had obesity rates of 20% by age 11y, four times the rate of similarly aged children twenty years prior [42].

Thus, in this study, we used data from the four British birth cohort studies to investigate how genetic penetrance upon childhood and adulthood BMI has changed over the obesity epidemic in the UK. We compared genotype-phenotype associations for two PGI derived from GWAS of adulthood and childhood BMI, respectively, as well as using genome-wide (i.e., SNP-heritability) and single gene (e.g., FTO) approaches. We further examined whether changes in the magnitudes of genetic effects have occurred across the entire distribution of population BMI. We hypothesised that changes in genetic penetrance would track cross-cohort differences in BMI: genetic effects would be stronger, at a given age, in each successive cohort, driven by stronger associations at upper centiles of the BMI distribution.

## Methods

### Participants

The MRC National Survey of Health and Development (hereafter, 1946c) follows a sample of individuals born in mainland Britain (England, Scotland, or Wales) during a single week of March 1946. Cohort members were recruited by sampling singleton births, with individuals from non-manual households oversampled. The National Child Development Study (hereafter, 1958c) and the British Cohort Study (hereafter, 1970c) track all individuals born in mainland Britain in single weeks of March 1958 and April 1970, respectively. Immigrants to the UK born in these weeks were later added to 1958c and 1970c using school enrolment information. The Millennium Cohort Study (hereafter, 2001c) follows a sample of individuals from across the UK born between 2000/02. Participants were recruited using a two-stage stratified sampling design and sampled from selected postcode areas. Individuals from Northern Ireland, Scotland and Wales, ethnic minority backgrounds, or disadvantaged areas were oversampled. Given differences in cohort eligibility and increasing ethnic diversity within the UK, we restricted our analysis in each cohort to singletons of White ethnicity born in England, Scotland, or Wales.

The 1946c, 1958c and 1970c were genotyped using whole blood samples collected at ages 53y, 44y, and 46y, respectively, while the 2001c were genotyped using saliva samples collected at 14y. The procedures used to genotype participants, as well as steps used to quality control (QC) and impute genetic data, are described further in the Supplementary Information. Procedures differed between each cohort. 2,731 (50.9%) eligible participants in the 1946c had genetic data. Corresponding figures were 5,989 (37.0%) for the 1958c, 5,170 (31.5%) for the 1970c and 5,489 (41.1%) for the 2001c.

Each cohort and survey sweep has received ethical approval and obtained appropriate consent according to guidance in place at data collection. Further details on each study are available in cohort profiles [43–48].

### Measures

#### Body Mass Index

Height and weight were obtained at the following ages:

- 1946c: ages 4y, 6y, 7y, 11y, 15y, 20y, 26y, 36y, 43y, 53y, 63y, and 69y
- 1958c: ages 7y, 11y, 16y, 23y, 33y, 42y, 44y, 50y, and 55y
- 1970c: ages 10y, 16y, 26y, 29y, 34y, 42y, and 46y
- 2001c: ages 3y, 5y, 7y, 11y, 14y and 17y

Height and weight were collected via direct measurement by interviewers, health visitors, doctors, or nurses except in the following sweeps where self-report was used: ages 20y and 26y in the 1946c; ages 23y, 42y, 50y and 55y in the 1958c; and ages 26y to 42y in the 1970c. Self-report was additionally used in a small number of cases where it was not possible to obtain a valid measurement from participants (e.g., where the participant refused).

We converted height and weight to BMI using standard formula (kg/m^2^). We excluded outlier values beyond +/- 3 SD of the sample mean (calculated in each cohort follow-up, separately), and from age 20+ used previous or succeeding measurements of adult height, where missing.

#### Polygenic Indices for Body Mass Index

In main analyses, we used two polygenic indices (PGIs) for adult BMI and child BMI, respectively, derived from genome-wide association studies (GWAS) of UK Biobank (UKB) data, a sample of approximately 500,000 British adults aged 39-73 year old at recruitment in 2006 -2010 [49]; the use of UKB avoided sample overlap with our data. Adulthood BMI was measured objectively at baseline assessment in UKB [50], while childhood weight was captured by retrospective self-report with participants asked whether at age ten they were “thinner, plumper, or about average” relative to others [51]. Previous work using the 1946c shows this PGI relates similarly to BMI as a PGI derived from a GWAS of prospectively measured child-adolescent BMI, but which used the 1958c in its discovery sample [52].

We calculated each PGI using PRSice-2 [53] limiting to clumped genome-wide significant hits (p < 5e-8, R^2^ < 0.01, 1,000 kb window) and disregarding ambiguous alleles, assuming additive genetic effects and, for comparability between cohorts, subsetting to single nucleotide polymorphisms (SNPs) genotyped or imputed in each cohort. For interpretability, we standardised the PGIs to have a mean of zero and a standard deviation of one across the combined sample. The final PGI scores were based on 505 (adulthood BMI) and 227 (childhood BMI) SNPs, respectively.

As BMI does not distinguish between fat and lean mass and height has increased over time [54], in sensitivity analysis, we alternatively used a PGI for adult fat mass percentage, specifically, again based on a GWAS of UKB data [50; 461 SNPs]. We additionally used a variable capturing the count of effect alleles for a variant within the *FTO* gene (rs1558902) that is related to fat mass and eating behaviour [55, 56].

#### Covariates and Auxiliary Variables

We included several variables as covariates. Depending on the model, these were the cohort member’s sex, verbal reasoning ability at age 10/11, maternal age at birth, mother’s years of education, family socioeconomic class, mother’s BMI, and cohort member’s first ten genetic principal components (PCs). Further detail on these variables is provided in the Supplementary Information.

### Statistical Analysis

To investigate changes in polygenic penetrance on (mean) BMI across cohorts, we regressed BMI values at each age upon the PGI, repeated separately for each PGI and cohort. As BMI was measured repeatedly, we used mixed effects modelling, with person-specific random intercepts added to models (observations nested within individuals). Given previous evidence that associations between PGI and BMI vary non-linearly over the life course [38], we interacted the PGI and age, with age modelled with two natural splines [57]. In our primary analysis, we adjusted for sex, a dummy variable for BMI measurement type (direct or self-report), and the first ten genetic principal components (PCs), the latter to account for population stratification [58]. From these regressions, we calculated marginal effects across the range follow-ups in a given cohort (i.e., ages 3y, 4y, …,17y in the 2001c) and then, for a given age in a pair of cohorts, calculated z-scores for differences in these marginal effects. We repeated each analysis, including additional adjustment for (a) family socioeconomic class, mother’s years of education and childhood cognitive ability and (b) mother’s age and BMI. Each of these were regarded as background factors that may explain changing associations.

To examine changes in the polygenic penetrance on the distribution of BMI across cohorts, we used quantile regression, estimating separate quantile regressions for each decile of BMI (10^th^, 20^th^, …, 90^th^ centiles), repeated for each PGI, age of follow-up, and cohort, and adjusting for sex, self-report dummy, (linear) age and the first ten genetic PCs. Estimates were then plotted and visually compared for each cohort, age, and outcome decile.

We performed two separate analyses to explore changes in the heritability of BMI across cohorts. First, we estimated ‘PGI-heritability’ by calculating incremental variance explained by each PGI. This was calculated by extracting R^2^ values from OLS regressions of BMI upon the PGI plus covariates (age, sex, and 10 PCs) and comparing these against R^2^ values obtained when not adjusting for the PGI. We repeated this for each PGI, cohort, and follow-up age separately and estimated 95% confidence intervals using bootstrapping (500 bootstraps, percentile method).

Second, we estimated SNP-heritability (SNP-h^2^) at each follow-up using Genomic Relatedness Restricted Maximum Likelihood (GREML), as implemented in the software GCTA [59]. This method exploits variation in genetic relatedness between sampled, not closely related individuals to calculate the proportion of phenotypic variance that can be explained (additively) by measured genetic variants [60]. The benefit of this approach is it does not rely on PGI weights, which here were based on the GWAS of an older population (i.e., UKB) and could, if causal genetic signals differ by cohort, be biased for younger cohorts.

We carried out all regression analyses using R version 4.3.1 [61]. Given the 1946c and 2001c used stratified study designs, we used study-specific probability (recruitment) weights in analyses, except for the GREML analysis, the software for which did not allow for the inclusion of weights. We used (regression-specific) complete case data. Sample sizes therefore differed across analyses due to missing data for PGI, BMI or covariates, loss to follow-up, death, and emigration. In sensitivity analyses, we created bespoke inverse probability weights to account for selection into the genotyped sample and combined this with multiple imputation to address remaining item missingness. The procedure we used is described further in the Supplementary Information.

## Results

### Descriptive Statistics

The mean and variance of BMI were higher at a given age in each successive cohort, though differences between the 1946c, 1958c, and 1970c only arose during early adulthood (Figure 1). The increase in variance was driven by increases at higher centiles of the BMI distribution – there was little difference between cohorts in the prevalence of underweight or median BMI, while differences at the 90^th^ centile were substantial. Mean BMI increased as each cohort aged, but the rate of increase was greater in younger cohorts.

**Figure 1:**
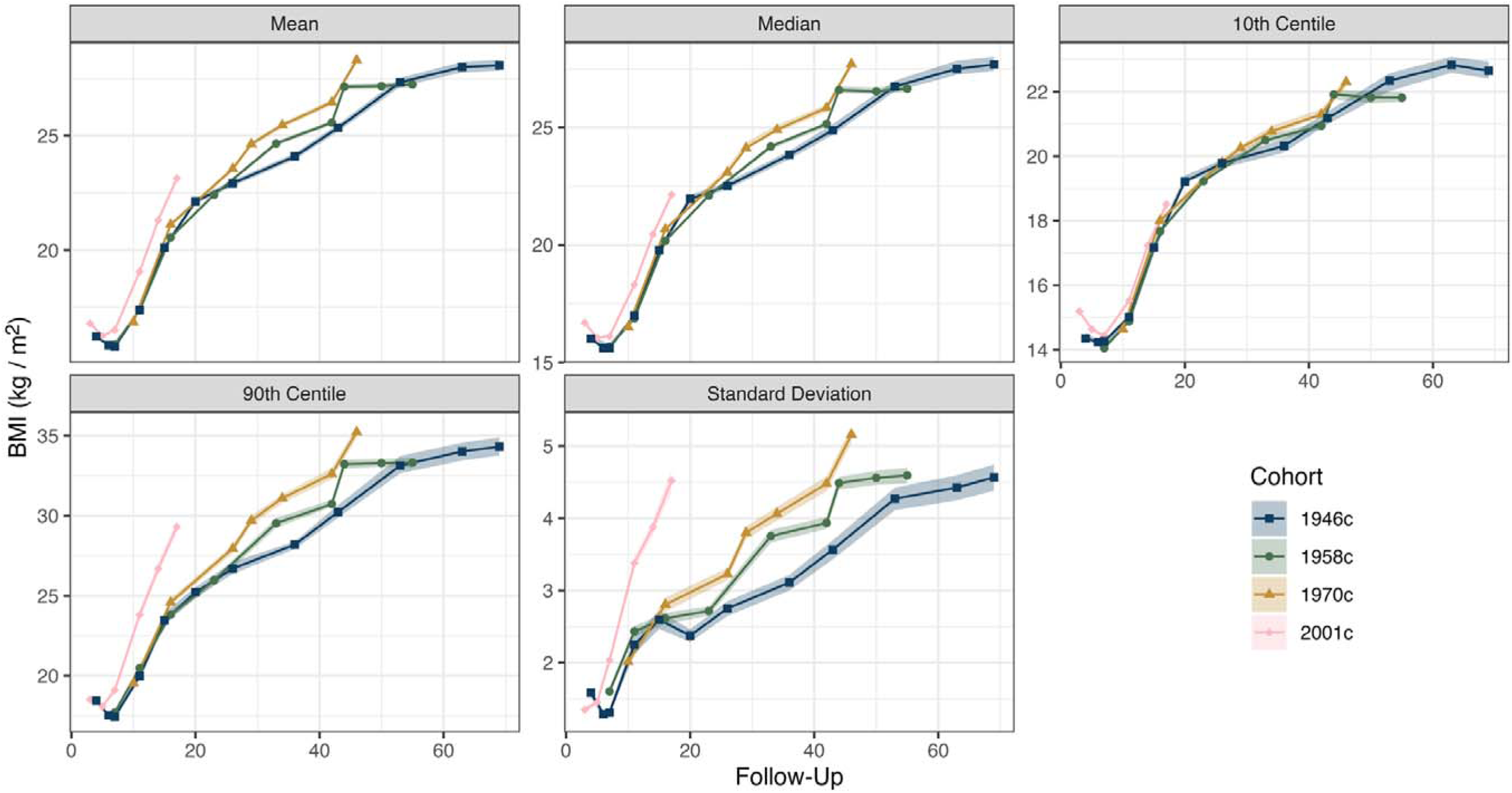
Descriptive statistics (+ 95% CIs) for BMI (kg/m^2^) by cohort and age at follow-up among genotyped participants. Estimates are weighted using recruitment weights and account for complex survey design.

PGI distributions were similar in each cohort (Supplementary Figure S1). Adulthood and childhood PGIs were positively correlated in each cohort; correlations ranged 0.34 – 0.36. The adulthood PGI was related to several covariates (Table 1), including positive correlations with mother’s (ρ = 0.08 – 0.14) and father’s BMI (ρ = 0.06 – 0.10) and negative correlations with verbal cognitive ability scores (ρ = -0.07 – -0.04). The adulthood PGI was also (negatively) related to parents’ education and maternal age. The childhood PGI was related to the mother’s and father’s BMI (ρ = 0.08 – 0.12 and 0.05 – 0.09, respectively; Supplementary Table S1). Associations with other covariates, including verbal cognitive ability scores, were weaker in size and close to the null.

**Table 1:**
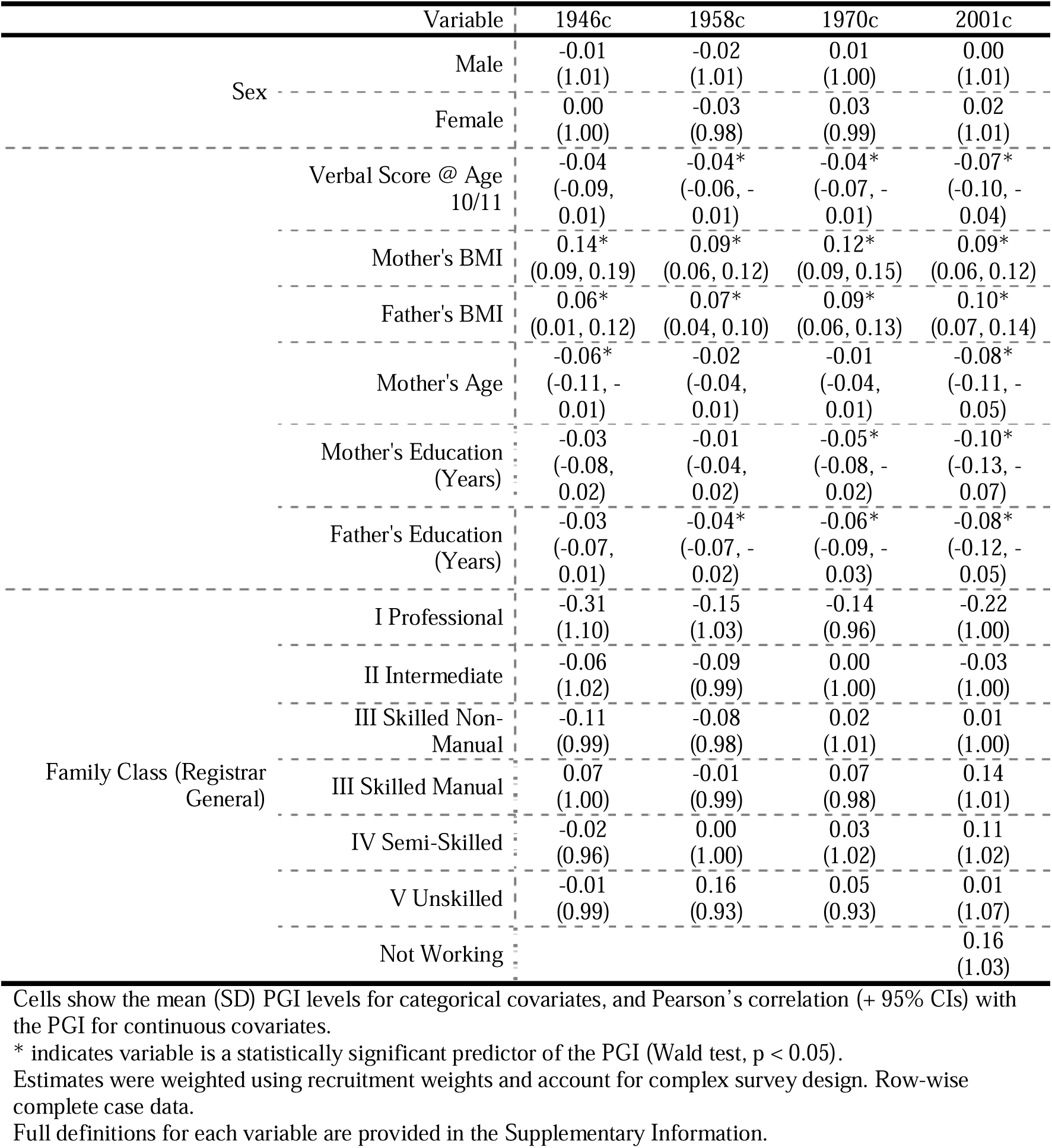
Association between adulthood PGI and covariates

There was evidence of selection bias in the genotyped samples. In each cohort, compared with other cohort members, genotyped individuals were more likely to be from advantaged socioeconomic backgrounds (as measured by family socioeconomic class and parental education) and had higher cognitive ability, on average (Supplementary Table S2). Higher BMI and PGI values were related to a greater likelihood of dropping out of each survey in later sweeps (Supplementary Figures S2 and S3).

### Changing Polygenic Penetrance on (Mean) BMI

The adult PGI was positively associated with BMI in each cohort in childhood, adolescence and adulthood (left panel, Figure 2). Associations strengthened as participants aged. These associations were stronger in successively younger cohorts – especially the 2001c – but the age at which differences between cohorts appeared grew earlier over time (top panels, Supplementary Figure S4).

**Figure 2:**
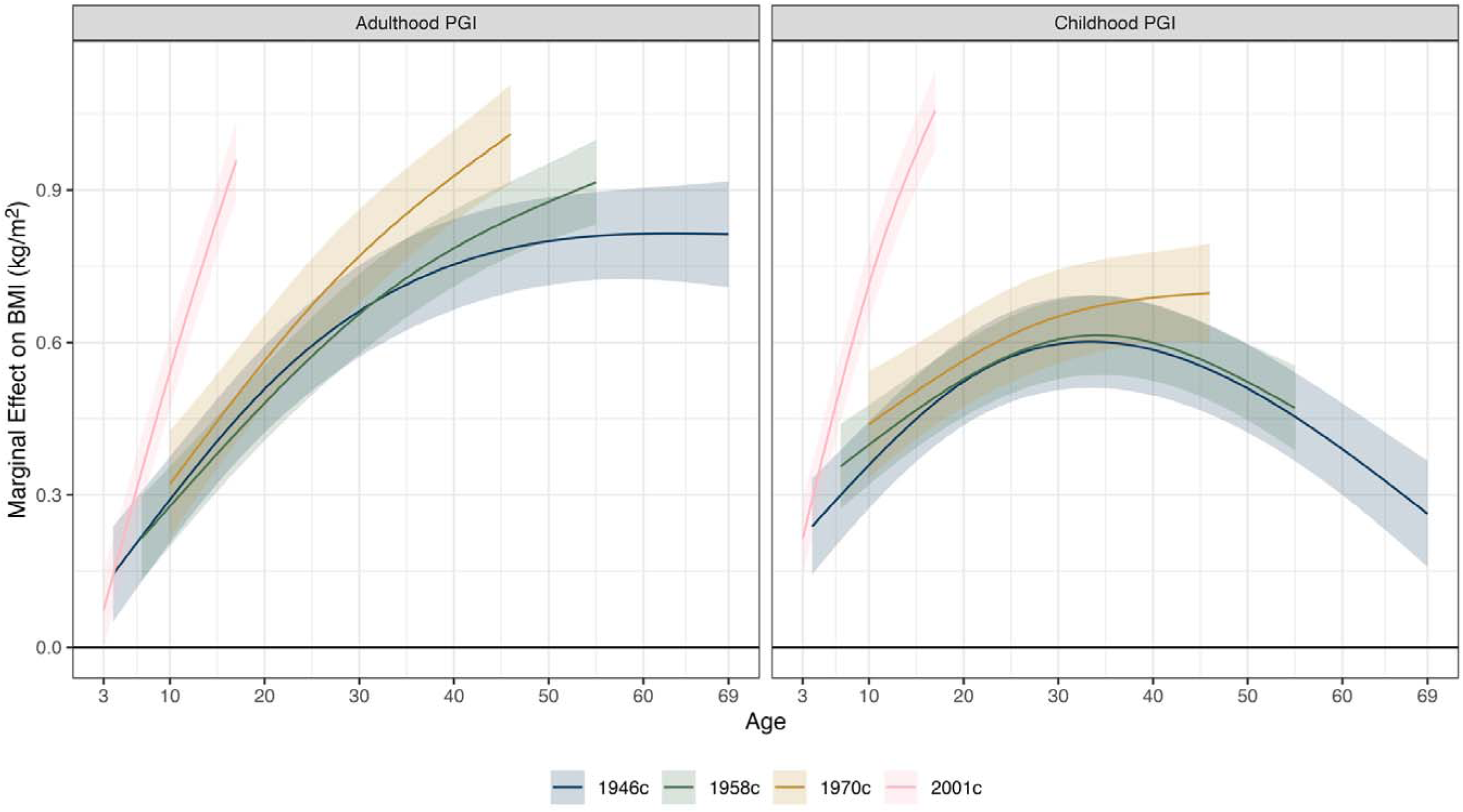
Association between PGI and BMI (kg/m^2^) by cohort, age, and PGI (adulthood or childhood). Derived from separate linear mixed effects models with association between PGI and BMI allowed to vary by age (two natural splines). Adjustment for age (two natural splines), sex, first 10 genetic principal components, and a person-specific random intercept. Estimates were weighted using recruitment weights.

For instance, associations between the adulthood PGI and BMI were consistently stronger in the 1958c than the 1946c only in mid-adulthood (∼ age 40+). In comparison, differences between the 2001c and earlier cohorts appeared from childhood. At age 16y, a 1 SD increase in the PGI was associated with a 0.90 kg/m^2^ (0.83, 0.97) higher BMI in the 2001c, over double the 0.43 kg/m^2^ (0.34, 0.51) difference estimated in the 1946c. Corresponding figures for the 1958c and 1970c were 0.40 kg/m^2^ (0.33, 0.48) and 0.47 kg/m^2^ (0.38, 0.56), respectively. At age 42y, a 1 SD increase in the PGI was associated with a higher BMI of 0.77 kg/m^2^ (0.68, 0.85) in the 1946c, 0.81 kg/m^2^ (0.73, 0.88) higher BMI in the 1958c, and a 0.96 kg/m^2^ (0.86, 1.05) higher BMI in the 1970c. Note, these are of comparable size to the associations as early as age 16y in the 2001c (for contour plot, see Supplementary Figure S5).

The childhood PGI was also positively associated with BMI in each cohort in childhood, adolescence and adulthood (right panel, Figure 2). Associations had an inverted-U shaped relationship with age, growing stronger into mid-adulthood but weaker thereafter. Associations were again stronger in successively younger cohorts, and notably larger in the 2001c (bottom panels, Supplementary Figure S4, and Supplementary Figure S6). However, there was little consistent difference between the 1946c and 1958c. At age 16y, a 1 SD increase in the PGI was associated with a higher BMI of 0.47 kg/m^2^ (0.38, 0.55) in the 1946c, 0.48 kg/m^2^ (0.41, 0.55) in the 1958c, 0.52 kg/m^2^ (0.42, 0.61) in the 1970c, and 1.01 kg/m^2^ (0.94, 1.09) in the 2001c.

### Changing Polygenic Penetrance on the Distribution of BMI

The association between the adulthood PGI and BMI was stronger at higher centiles of BMI in each cohort, indicating greater variance and skewness in BMI among those with higher PGI values (selected results shown in Figure 3; full results shown in Supplementary Figures S7-S8). Differences between the 2001c and the earlier cohorts in the association between the adulthood PGI and BMI were more pronounced at higher centiles of the distribution. At age 10/11, at the 10^th^ centile of the BMI distribution, a 1 SD increase in the adulthood PGI was associated with 0.17 kg/m^2^ (0.06, 0.28) higher BMI in the 1946c, 0.10 kg/m^2^ (0.06, 0.15) in the 1958c, 0.18 kg/m^2^ (0.11, 0.25) in the 1970c and 0.25 kg/m^2^ (0.18, 0.31) in the 2001c. Corresponding figures at the same age for the 90^th^ centile were 0.62 kg/m^2^ (0.36, 0.88), 0.77 kg/m^2^ (0.61, 0.92), 0.50 kg/m^2^ (0.34, 0.66),and 1.20 kg/m^2^ (0.99, 1.40), respectively. Differences in association in adulthood between the oldest three cohorts were less pronounced. Stronger associations at higher centiles, particularly for the 2001c, were generally also observed when examining the association between the childhood PGI and BMI scores (Supplementary Figures S8-S9).

**Figure 3:**
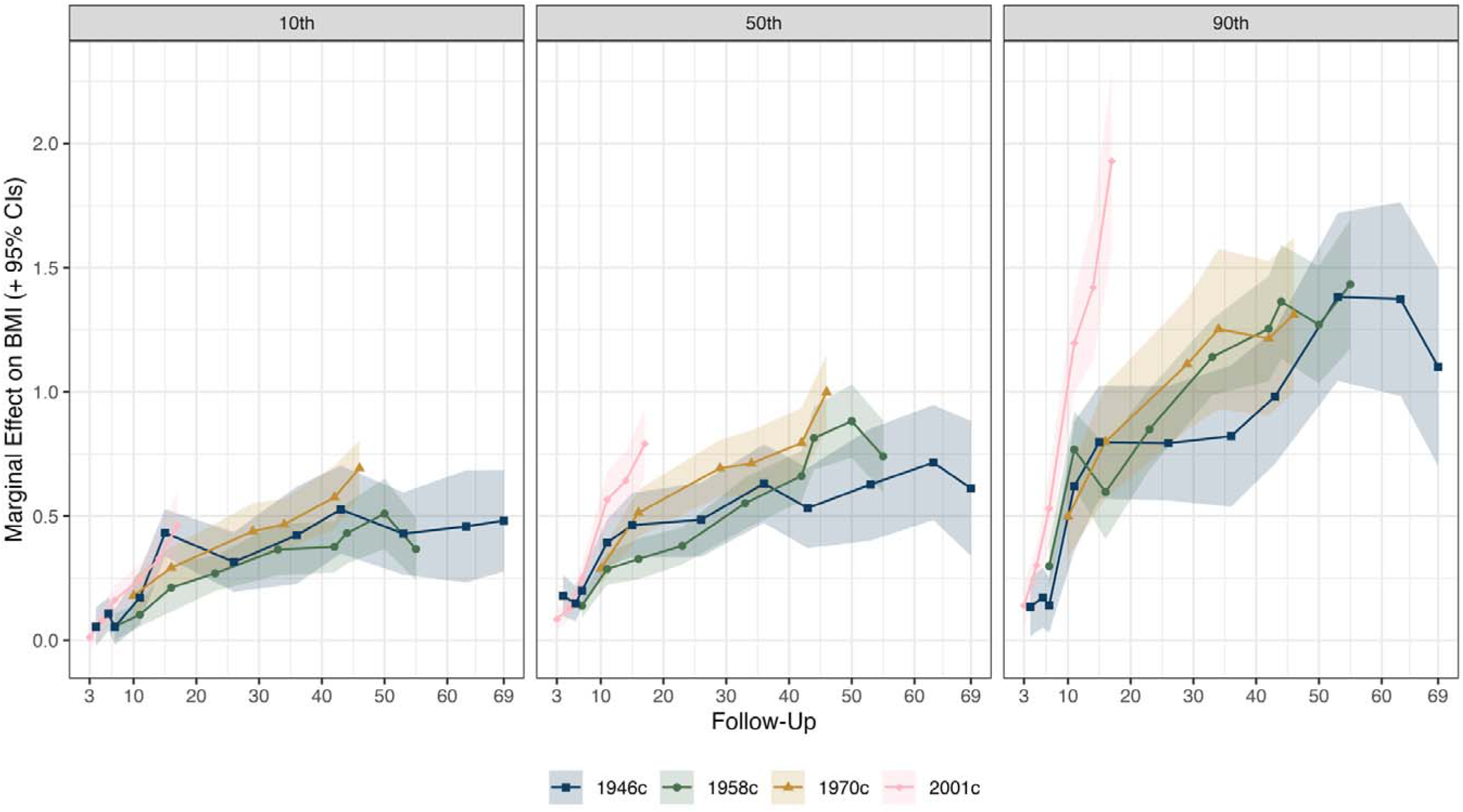
Association between adulthood PGI and BMI (kg/m^2^) by BMI decile, cohort, age of follow-up. Derived from separate quantile regressions adjusting for age (linear term), sex and first 10 genetic principal components. Estimates were weighted using recruitment weights. Each panels displays associations for a particular, selected decile of BMI (10^th^, 50^th^, 90^th^). Results show how the (conditional) centiles of BMI vary according to 1 SD increases in the adulthood PGI. Full results (10^th^, 20^th^, …, 90^th^ deciles) are displayed in Supplementary Figure S6.

### Changes in the Heritability of BMI

Though associations between the adulthood PGI and BMI increased as participants aged (top left panel, Figure 4), the variance in BMI explained by the adulthood PGI (i.e., PGI-heritability) stayed largely constant across adulthood (middle left panel, Figure 4; also see Supplementary Table S3) reflecting the increasing variance in BMI at older ages. The adulthood PGI explained at most 4.1% of the variance in BMI in the 1946c (43y; 95% CI = 2.5%, 5.9%), 3.7% in the 1958c (50y; 95% CI = 2.7%, 4.6%), 3.9% in the 1970c (29y; 95% CI = 2.9%, 5.0%), and 4.3% in the 2001c (17y; 95% CI = 3.2%, 5.5%). There were no clear, consistent cohort differences in PGI-heritability using the adulthood PGI.

**Figure 4:**
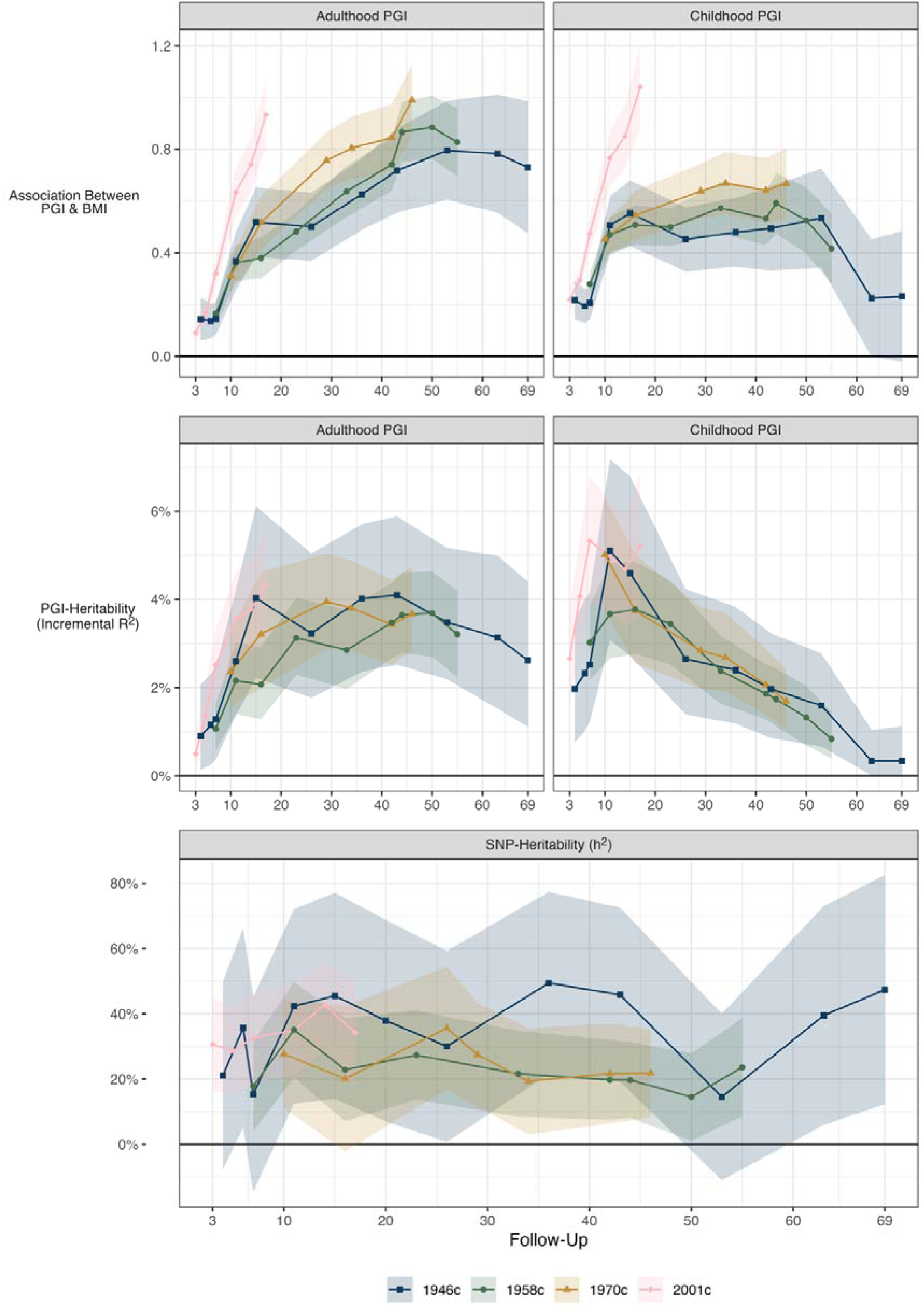
Heritability of BMI and association between adulthood and childhood PGI and BMI by cohort and follow-up. Top Panels: Regression of BMI (kg/m^2^) upon PGI by cohort, age of follow-up, and PGI (adulthood or childhood). Derived from separate OLS regressions adjusting for age, sex and first 10 genetic principal components. Estimates were weighted using recruitment weights and account for complex survey design. Marginal effect of 1 SD higher PGI on BMI (kg/m^2^). Middle Panels: incremental proportion of variance explained by (adulthood or childhood) PGI calculated by comparing R^2^ with regression of BMI on age, sex and first 10 genetic principal components, with and without further adjustment for the PGI. Confidence intervals estimated using bootstrapping (500 bootstraps, percentile method). Bottom Panel: SNP-heritability of BMI calculated with GCTA, adjusting for sex, age and first 10 genetic principal components. Survey weights were not incorporated in the GCTA analysis.

The proportion of variance in BMI explained by the childhood PGI increased into adolescence but declined thereafter (middle right panel, Figure 4; also see Supplementary Table S3). The childhood PGI explained at most 5.1% of the variance in BMI in the 1946c (11y; 95% CI = 3.1%, 7.2%), 3.8% in the 1958c (16y; 95% CI = 2.8%, 4.9%), 5.0% in the 1970c (10y; 95% CI = 3.7%, 6.3%), and 5.3% in the 2001c (7y; 95% CI = 4.0%, 6.8%) Again, there were no clear and consistent cohort differences in PGI-heritability using the childhood PGI.

SNP-h^2^ estimates estimated with GREML were larger than PGI-heritability estimates and ranged 14.5% (53y) to 49.5% (36y) in the 1946c, 14.5% (50y) to 35.1% (11y) in the 1958c, 19.3% (34y) to 35.6% (26y) in the 1970c, and 28.6% (5y) to 42.7% (14y) in the 2001c (bottom panel, Figure 4; also see Table S3). SNP-h^2^ was greatest in the 1946c, but at each follow-up and in each cohort, confidence intervals were wide. Given the lack of precision in the estimates, there was no clear trend in SNP-h^2^ by age.

### Sensitivity Analyses

Results for the associations between each PGI and mean BMI (kg/m^2^) were qualitatively similar when additionally controlling for family socioeconomic position and childhood cognitive ability or maternal BMI and maternal age at birth (Supplementary Figure S10). Results were also qualitatively similar when using weighting to account for selection into the genotyped sample, though cohort differences in later adulthood were less pronounced (Supplementary Figure S11). Mixed effects model results were partly replicated when using the PGI for adult fat mass rather than a PGI for adult BMI, specifically: the PGI had the strongest association in the youngest cohort (bottom left panel, Supplementary Figure S12). This was also the case when using the rs1558902 *FTO* variant, though confidence intervals overlapped owing to the lower predictive power of the variable (bottom right panel, Supplementary Figure S12).

## Discussion

### Summary of Results

Using multiple national birth cohorts with data spanning 1950 to 2018, we found considerable cross-cohort differences in associations of genetics with BMI. In each cohort, the adulthood PGI-BMI association increased as individuals aged, but the relationship was strongest in the most recent-born cohort (2001c). Differences in the strength of association of the adulthood PGI between the 1946c, 1958c and 1970c arose during adulthood, tracking the development of cross-cohort differences in BMI among these cohorts. Associations between the adulthood PGI and BMI were stronger at higher centiles of BMI in each cohort, consistent with genetic effects disproportionately impacting obesity rates. Cross-cohort differences in the effects were also typically larger at higher centiles of the BMI distribution. Again, this tracked the nature of cross-cohort differences in BMI which were driven by increases in obesity, in particular. Cross-cohort differences were also observed using PGI for childhood BMI and adult fat mass.

### Explanation of Results

The finding of larger associations between PGI and BMI in cohorts more affected by the obesity epidemic is consistent with previous studies in adults and twins [26–36, 62]. We extend these results by examining data across life in multiple national cohorts and undertaking a range of analyses from distributional modelling to PGI, genome-wide, and specific gene approaches to inference. Changes in genetic penetrance appear to have tracked the obesity epidemic – specifically, the timing at which differences in phenotypic BMI across cohorts has arisen and the disproportionate effects on obesity rates.

Why the adulthood and childhood PGIs have stronger associations with BMI in more recent born cohorts is unclear. While birth year is arguably an exogenous source of environmental variation, it does not distinguish which aspects of the environment have led to the changes we observed. Previous studies have shown weaker effects of PGI on BMI among physically active individuals [63] and those living in deprived neighbourhoods [64] or in greater proximity to fast-food restaurants [65, though also see 66]. As the environment has changed, it may have enabled greater expression of genetic liability towards higher calorie consumption and, thus, higher BMI.

Factors that predict increasing genetic penetrance over time may also explain the stronger effects we observed at the upper centiles of the BMI distribution. In the 2001c, for example, proximity to fast food outlets varies considerably between cohort members and across time [67]. If genetic effects are of greater importance where calorific food is more readily available, we would, therefore, expect heterogeneous genetic effects. This would be reflected as greater variation and skewness in the distribution of BMI over time, as observed here. Further, some, but not all, individuals may offset greater genetic risk through higher physical activity or other changes in behaviour [63], which may again lead to greater variation in BMI among those with high PGI values.

Cohort differences may also be driven by genetically-influenced nurture. Recent work suggests that maternal BMI-related genetic variants that are not directly inherited influence offspring BMI [68–70]. Parents make choices on children’s behalf, and children may also model parent behaviour. Given changes in adult eating and exercise habits over time, this could explain some of the change in genetic effects in the 2001c relative to earlier cohorts. An extension of this work would be to investigate how much of the effects on mean and variance are due to direct versus indirect genetic effects.

### Strengths and Limitations

Strengths include using BMI data collected from nationally representative samples at multiple ages in each cohort, particularly measurements from early childhood and adolescence. BMI was also measured objectively on most measurement occasions and the cohorts we used spanned a wide period of recent history, including before the obesity epidemic in the UK. Data collection also overlapped with a period of post-war food rationing for the 1946c.

Limitations include the high degree of attrition (>50%) reflected in the genotyped samples. Individuals with higher BMI had higher rates of drop-out in each cohort, which may have biased results. However, similar results were obtained when accounting for selection into the genotyped sample with inverse probability weighting. Our study relied upon GWAS of an older cohort that did not span all age ranges or birth years used here. However, arguably this should bias towards finding larger effects in older cohorts, the opposite of what we found here. While the procedures used to QC and impute genetic data were harmonised across each cohort, different genotypic chips were used, which may have biased results.

### Conclusions

Identical genetic variation appears to have had more pronounced consequences for BMI in cohorts born later in the obesity epidemic. Genetic associations were stronger at highest BMI centiles – the part of the distribution that has changed most in recent decades. Findings suggest that the effects of genes on BMI are, to some extent, modifiable. Future research should identify aspects of the environment that can temper genetic predisposition.

## Data Availability

The code used to run the analysis will be made available at https://osf.io/nxusz. Genetic data for the Millennium Cohort Study, British Cohort Study, National Child Development Study, and National Survey of Health and Development are available via application from the respective data owners. Summary statistics from the GWAS of adulthood BMI and adulthood fat mass were downloaded from the Neale Lab website (http://www.nealelab.is/uk-biobank; UKB data-fields 23104 and 23099). Summary statistics from the GWAS of (recalled) childhood BMI were downloaded from the PGS Catalog (https://www.pgscatalog.org/score/PGS000716).

## References

1. Dai H, Alsalhe TA, Chalghaf N, Riccò M, Bragazzi NL, Wu J. The global burden of disease attributable to high body mass index in 195 countries and territories, 1990–2017: An analysis of the Global Burden of Disease Study. PLoS Med. 2020;17:e1003198.

2. World Obesity Federation. World Obesity Atlas 2023. London; 2023.

3. Baker C. Obesity statistics. Research Briefing. House of Commons Library; 2023.

4. Norris T, Bann D, Hardy R, Johnson W. Socioeconomic inequalities in childhood-to-adulthood BMI tracking in three British birth cohorts. Int J Obes. 2020;44:388–98.

5. Johnson W, Li L, Kuh D, Hardy R. How Has the Age-Related Process of Overweight or Obesity Development Changed over Time? Co-ordinated Analyses of Individual Participant Data from Five United Kingdom Birth Cohorts. PLoS Med. 2015;12:e1001828.

6. Park MH, Falconer C, Viner RM, Kinra S. The impact of childhood obesity on morbidity and mortality in adulthood: a systematic review. Obesity Reviews. 2012;13:985–1000.

7. Abarca-Gómez L, Abdeen ZA, Hamid ZA, Abu-Rmeileh NM, Acosta-Cazares B, Acuin C, et al. Worldwide trends in body-mass index, underweight, overweight, and obesity from 1975 to 2016: a pooled analysis of 2416 population-based measurement studies in 128·9 million children, adolescents, and adults. The Lancet. 2017;390:2627–42.

8. Green MA, Subramanian SV, Razak F. Population-level trends in the distribution of body mass index in England, 1992–2013. J Epidemiol Community Health. 2016;70:832–5.

9. Office for Health Improvement and Disparities. Child obesity: patterns and trends. 2022.

10. Prentice AM, Jebb SA. Obesity in Britain: gluttony or sloth? BMJ. 1995;311:437–9.

11. Stamatakis E, Primatesta P, Chinn S, Rona R, Falascheti E. Overweight and obesity trends from 1974 to 2003 in English children: what is the role of socioeconomic factors? Archives of Disease in Childhood. 2005;90:999–1004.

12. Cutler DM, Glaeser EL, Shapiro JM. Why Have Americans Become More Obese? Journal of Economic Perspectives. 2003;17:93–118.

13. Keith SW, Redden DT, Katzmarzyk PT, Boggiano MM, Hanlon EC, Benca RM, et al. Putative contributors to the secular increase in obesity: exploring the roads less traveled. Int J Obes (Lond). 2006;30:1585–94.

14. McAllister EJ, Dhurandhar NV, Keith SW, Aronne LJ, Barger J, Baskin M, et al. Ten Putative Contributors to the Obesity Epidemic. Critical Reviews in Food Science and Nutrition. 2009;49:868– 913.

15. Millward DJ. Energy balance and obesity: a UK perspective on the gluttony v. sloth debate. Nutrition Research Reviews. 2013;26:89–109.

16. Government Office for Science. Tackling Obesities: Future Choices – Project Report. 2007.

17. Fox KR, Hillsdon M. Physical activity and obesity. Obesity Reviews. 2007;8:115–21.

18. Griffith R, Jin W (Michelle), Lechene V. The decline of home-cooked food. Fiscal Studies. 2022;43:105–20.

19. Jones P. The growth of fast food operations in Britain. Geography. 1985;70:347–50.

20. Flegal KM, Carroll MD, Kit BK, Ogden CL. Prevalence of Obesity and Trends in the Distribution of Body Mass Index Among US Adults, 1999-2010. JAMA. 2012;307:491–7.

21. Elks CE, den Hoed M, Zhao JH, Sharp SJ, Wareham NJ, Loos RJF, et al. Variability in the heritability of body mass index: a systematic review and meta-regression. Front Endocrinol (Lausanne). 2012;3:29.

22. Jackson SE, Llewellyn CH, Smith L. The obesity epidemic – Nature via nurture: A narrative review of high-income countries. SAGE Open Medicine. 2020;8:2050312120918265.

23. Frayling TM, Timpson NJ, Weedon MN, Zeggini E, Freathy RM, Lindgren CM, et al. A common variant in the FTO gene is associated with body mass index and predisposes to childhood and adult obesity. Science. 2007;316:889–94.

24. Speakman JR. The ‘Fat Mass and Obesity Related’ (FTO) gene: Mechanisms of Impact on Obesity and Energy Balance. Curr Obes Rep. 2015;4:73–91.

25. Sahoo K, Sahoo B, Choudhury A, Sofi N, Kumar R, Bhadoria A. Childhood obesity: causes and consequences. J Family Med Prim Care. 2015;4:187.

26. Brandkvist M, Bjørngaard JH, Ødegård RA, Åsvold BO, Sund ER, Vie GÅ. Quantifying the impact of genes on body mass index during the obesity epidemic: longitudinal findings from the HUNT Study. BMJ. 2019;:l4067.

27. Brandkvist M, Bjørngaard JH, Ødegård RA, Brumpton B, Smith GD, Åsvold BO, et al. Genetic associations with temporal shifts in obesity and severe obesity during the obesity epidemic in Norway: A longitudinal population-based cohort (the HUNT Study). PLOS Medicine. 2020;17:e1003452.

28. Conley D, Laidley TM, Boardman JD, Domingue BW. Changing Polygenic Penetrance on Phenotypes in the 20th Century Among Adults in the US Population. Sci Rep. 2016;6:30348.

29. Demerath EW, Choh AC, Johnson W, Curran JE, Lee M, Bellis C, et al. The Positive Association of Obesity Variants with Adulthood Adiposity Strengthens over an 80-Year Period: A Gene-by-Birth Year Interaction. Hum Hered. 2013;75:175–85.

30. Guo G, Liu H, Wang L, Shen H, Hu W. The Genome-Wide Influence on Human BMI Depends on Physical Activity, Life Course, and Historical Period. Demography. 2015;52:1651–70.

31. Liu H, Guo G. Lifetime Socioeconomic Status, Historical Context, and Genetic Inheritance in Shaping Body Mass in Middle and Late Adulthood. Am Sociol Rev. 2015;80:705–37.

32. Rosenquist JN, Lehrer SF, O’Malley AJ, Zaslavsky AM, Smoller JW, Christakis NA. Cohort of birth modifies the association between FTO genotype and BMI. Proc Natl Acad Sci USA. 2015;112:354–9.

33. Sarnowski C, Conomos MP, Vasan RS, Meigs JB, Dupuis J, Liu C-T, et al. Genetic Effect on Body Mass Index and Cardiovascular Disease Across Generations. Circ: Genomic and Precision Medicine. 2023;:e003858.

34. Walter S, Mejía-Guevara I, Estrada K, Liu SY, Glymour MM. Association of a Genetic Risk Score With Body Mass Index Across Different Birth Cohorts. JAMA. 2016;316:63.

35. Rokholm B, Silventoinen K, Tynelius P, Gamborg M, Sørensen TIA, Rasmussen F. Increasing Genetic Variance of Body Mass Index during the Swedish Obesity Epidemic. PLoS ONE. 2011;6:e27135.

36. Rokholm B, Silventoinen K, Ängquist L, Skytthe A, Kyvik KO, Sørensen TIA. Increased Genetic Variance of BMI with a Higher Prevalence of Obesity. PLoS ONE. 2011;6:e20816.

37. Sanz-de-Galdeano A, Terskaya A, Upegui A. Association of a genetic risk score with BMI along the life-cycle: Evidence from several US cohorts. PLoS ONE. 2020;15:e0239067.

38. Bann D, Wright L, Hardy R, Williams DM, Davies NM. Polygenic and socioeconomic risk for high body mass index: 69 years of follow-up across life. PLoS Genet. 2022;18:e1010233.

39. Pearson HW. The life project: the extraordinary story of our ordinary lives. London: Allen Lane, an imprint of Penguin Books; 2016.

40. Prynne CJ, Paul AA, Price GM, Day KC, Hilder WS, Wadsworth MEJ. Food and nutrient intake of a national sample of 4-year-old children in 1950: comparison with the 1990s. Public Health Nutrition. 1999;2:537–47.

41. Prynne CJ, Paul AA, Mishra GD, Hardy RJ, Bolton-Smith C, Wadsworth MEJ. Sociodemographic inequalities in the diet of young children in the 1946 British birth cohort. Public Health Nutrition. 2002;5:733–45.

42. Connelly R, Chatzitheochari S. Chapter 6: Physical Development. In: Platt L, editor. Millennium Cohort Study: Initial findings from the Age 11 survey. London: Centre for Longitudinal Studies; 2014. p. 65–76.

43. Connelly R, Platt L. Cohort Profile: UK Millennium Cohort Study (MCS). International Journal of Epidemiology. 2014;43:1719–25.

44. Elliott J, Shepherd P. Cohort Profile: 1970 British Birth Cohort (BCS70). International Journal of Epidemiology. 2006;35:836–43.

45. Kuh D, M P, J A, J D, U E, P F, et al. Updating the cohort profile for the MRC National Survey of Health and Development: a new clinic-based data collection for ageing research. International Journal of Epidemiology. 2011;40:e1–9.

46. Power C, Elliott J. Cohort profile: 1958 British birth cohort (National Child Development Study). International Journal of Epidemiology. 2006;35:34–41.

47. Sullivan A, Brown M, Hamer M, Ploubidis GB. Cohort Profile Update: The 1970 British Cohort Study (BCS70). Int J Epidemiol. 2023;52:e179–86.

48. Wadsworth M, Kuh D, Richards M, Hardy R. Cohort Profile: The 1946 National Birth Cohort (MRC National Survey of Health and Development). International Journal of Epidemiology. 2006;35:49–54.

49. Sudlow C, Gallacher J, Allen N, Beral V, Burton P, Danesh J, et al. UK Biobank: An Open Access Resource for Identifying the Causes of a Wide Range of Complex Diseases of Middle and Old Age. PLOS Medicine. 2015;12:e1001779.

50. Neale Lab. UK Biobank GWAS Results. 2018. http://www.nealelab.is/uk-biobank/ww.nealelab.is/uk-biobank/. Accessed 13 Apr 2024.

51. Richardson TG, Sanderson E, Elsworth B, Tilling K, Davey Smith G. Use of genetic variation to separate the effects of early and later life adiposity on disease risk: mendelian randomisation study. BMJ. 2020;:m1203.

52. Vogelezang S, Bradfield JP, Ahluwalia TS, Curtin JA, Lakka TA, Grarup N, et al. Novel loci for childhood body mass index and shared heritability with adult cardiometabolic traits. PLoS Genet. 2020;16:e1008718.

53. Choi SW, O’Reilly PF. PRSice-2: Polygenic Risk Score software for biobank-scale data. GigaScience. 2019;8:giz082.

54. Bann D, Wright L, Davies NM, Moulton V. Weakening of the cognition and height association from 1957 to 2018: Findings from four British birth cohort studies. eLife. 2023;12:e81099.

55. Micali N, Field AE, Treasure JL, Evans DM. Are obesity risk genes associated with binge eating in adolescence? Obesity. 2015;23:1729–36.

56. Berndt SI, Gustafsson S, Mägi R, Ganna A, Wheeler E, Feitosa MF, et al. Genome-wide meta-analysis identifies 11 new loci for anthropometric traits and provides insights into genetic architecture. Nat Genet. 2013;45:501–12.

57. Perperoglou A, Sauerbrei W, Abrahamowicz M, Schmid M. A review of spline function procedures in R. BMC Med Res Methodol. 2019;19:46.

58. McVean G. A Genealogical Interpretation of Principal Components Analysis. PLOS Genetics. 2009;5:e1000686.

59. Yang J, Lee SH, Goddard ME, Visscher PM. GCTA: A Tool for Genome-wide Complex Trait Analysis. Am J Hum Genet. 2011;88:76–82.

60. Barry C-JS, Walker VM, Cheesman R, Davey Smith G, Morris TT, Davies NM. How to estimate heritability: a guide for genetic epidemiologists. International Journal of Epidemiology. 2023;52:624– 32.

61. R Core Team. R: A Language and Environment for Statistical Computing. 2023.

62. Stutzmann F, Tan K, Vatin V, Dina C, Jouret B, Tichet J, et al. Prevalence of Melanocortin-4 Receptor Deficiency in Europeans and Their Age-Dependent Penetrance in Multigenerational Pedigrees. Diabetes. 2008;57:2511–8.

63. CelislJMorales CA, Lyall DM, Bailey MES, PetermannlJRocha F, Anderson J, Ward J, et al. The Combination of Physical Activity and Sedentary Behaviors Modifies the Genetic Predisposition to Obesity. Obesity. 2019;27:653–61.

64. Tyrrell J, Wood AR, Ames RM, Yaghootkar H, Beaumont RN, Jones SE, et al. Gene–obesogenic environment interactions in the UK Biobank study. International Journal of Epidemiology. 2017;46:559–75.

65. Mason KE, Palla L, Pearce N, Phelan J, Cummins S. Genetic risk of obesity as a modifier of associations between neighbourhood environment and body mass index: an observational study of 335 046 UK Biobank participants. BMJ Nutr Prev Health. 2020;3:247–55.

66. Burgoine T, Monsivais P, Sharp SJ, Forouhi NG, Wareham NJ. Independent and combined associations between fast-food outlet exposure and genetic risk for obesity: a population-based, cross-sectional study in the UK. BMC Medicine. 2021;19:49.

67. Libuy N, Church D, Ploubidis G, Fitzsimons E. Fast food proximity and weight gain in childhood and adolescence: Evidence from Great Britain. Health Economics. 2024;33:449–65.

68. Bond TA, Richmond RC, Karhunen V, Cuellar-Partida G, Borges MC, Zuber V, et al. Exploring the causal effect of maternal pregnancy adiposity on offspring adiposity: Mendelian randomisation using polygenic risk scores. BMC Med. 2022;20:34.

69. Tubbs JD, Porsch RM, Cherny SS, Sham PC. The Genes We Inherit and Those We Don’t: Maternal Genetic Nurture and Child BMI Trajectories. Behav Genet. 2020;50:310–9.

70. Wright L, Shireby G, Morris TT, Davies NM, Bann D. The Association Between Parental BMI and Offspring Adiposity: A Genetically Informed Analysis of Trios. 2024;:2024.03.07.24303912.

