## Supplementary Information for "Genetic Risk for High Body Mass Index Before and Amidst the Obesity Epidemic: Cross-Cohort Analysis of Four British Birth Cohort Studies"

### Methods

#### Genotyping

Cohorts were genotyped using different chips, but similar procedures were used in each cohort to quality control (QC) and impute the genetic data and we further restricted the genotype files to include only those SNPs in common across the four cohorts. Text on the QC and imputation of 1958c, 1970c and 2001c genetic data is taken from the Centre for Longitudinal Studies (2024) Genomics Data website.

### 1946c

The 1946c were genotyped using whole blood samples collected at 53y (Hardy et al., 2010). A total of 2,851 individuals were genotyped using the DrugDev microarray (assaying 476,728 SNPs) platform. Genotype calling was performed using GenomeStudio (v2.0, Illumina) and quality control was completed using PLINK 1.9 and 2.0. Samples were read into GenomeStudio (0-1.27% samples excluded) and mapped to a manifest file. Individuals were excluded if they had (i) >2% missing data (1.88%-3.50% samples excluded), (ii) their genotype predicted sex using X chromosome homozygosity was discordant with their reported sex (excluding females with an F value > 0.2 and males with an F value < 0.8) (0.18%-2.14% samples excluded) (iii) they had excess heterozygosity [> three standard deviations (SD) from the mean] (0.36%-1.07% samples excluded), (iv) For related individuals in 1946c, 1958c, and 1970c the King algorithm (king-cutoff 0.0884) was employed to identify and exclude one individual from each pair of closely related individuals (3rd degree or closer) (0.19%-0.59% samples excluded). Duplicate samples were removed, retaining those with the higher genotyping rate.

We identified European samples by (i) merging the genotypes with data from 1000 genomes Phase 3, (ii) linkage disequilibrium pruning the overlapping single nucleotide polymorphisms (SNPs) such that no pair of SNPs within 1000 bp had r2 > 0.20 and (iii) using an elastic net model to establish which of the super populations the samples fall into (Africans [AFR], Admixed Americans [AMR], East Asians [EAS], Europeans [EUR] and South Asians [SAS]). Although this method puts each sample into the nearest superpopulation, there are still ancestral outliers. We advise these are removed based on principal components. We retain samples from all ancestry and provide a variable to capture this. Before imputation, SNPs with high levels of missing data (>3%), Hardy-Weinberg equilibrium P<1e-6 or minor allele frequency <1% were excluded.

The genetic data were then recoded as .vcf files before uploading to the TOPMed Imputation Server which uses Eagle2 to phase haplotypes, and Minimac4 (<https://genome.sph.umich.edu/wiki/Minimac4>) with the TOPMed reference panel. The genome build was updated to hg38 using LiftOver, implemented within the TOPMed server. Imputed genotypes were then filtered with PLINK2.0alpha, excluding SNPs with an R2 INFO score < 0.8 and recoded as binary PLINK format. Proceeding with PLINK1.9, samples with >2% missing values, SNPs with >2 alleles, >3% missing values, Hardy-Weinberg equilibrium P<1e-6 or a minor allele frequency of <1% were excluded (indels have not been excluded).

2,731 (50.9%) eligible participants in the 1946c had (valid) genetic data.

### 1958c

The 1958c were genotyped using whole blood samples collected at 44y (Bridges et al., 2023). Genotyping for 13,738 samples (6,431 unique individuals) was performed across seven different genotyping arrays. Quality control was completed using PLINK1.9, PLINK2.0, R v3.3.2 and RStudio v4.1.2. Each dataset was updated to GRCh37 build for consistency, using up-to-date strand files and a series of commands collated in the script Update Build (Robertson, 2012) or using the LiftOver software tool. For each chip individuals were excluded if they had (i) they had > 2% missing data (ii) their genotype predicted sex using X chromosome homozygosity was discordant with their reported sex (excluding females with an F value > 0.2 and males with an F value < 0.8) (iii) they had excess heterozygosity [>3 standard deviation (SD) from the mean] and (iv) they were related to another individual in the sample (–genome threshold 0.1875), removing samples with the most missing data. Prior to imputation SNPs with high levels of missing data (>3%), Hardy-Weinberg equilibrium P < 1x10-6 or minor allele frequency <1% were excluded. The genetic data were then recoded as vcf files before uploading to the Michigan Imputation Server which uses Eagle2 to phase haplotypes, and Minimac4 (<https://genome.sph.umich.edu/wiki/Minimac4>) with the HRC r1.1 reference panel.

Imputed genotypes were then filtered with PLINK2.0alpha, excluding SNPs with an R2 INFO score < 0.8 and recoded as binary PLINK format. Proceeding with PLINK1.9, samples with >2% missing values, and SNPs with >2 alleles, >3% missing values, Hardy-Weinberg equilibrium P < 1e-6 or a minor allele frequency of <1% were excluded. We combined data from five of the seven chips (Illumina 1.2M, Illumina Human 660-Quad, Infinium HumanHap 550K v1.1, Infinium HumanHap 550K v3 and Affymetrix v6) which had, high and similar imputation quality (based on number of SNPs after QC). The Illumina 15k Custom Chip and Affymetrix 500k were not included in the combined dataset since they produced lower quality results, yielding less high-quality imputed SNPs than the other arrays. It is worth noting all samples, bar five, were covered by the other arrays and the combined dataset consisted of 6,420 individuals and 6,722,830 SNPs.

Further QC was conducted on the combined dataset where individuals were excluded if they had (i) they had > 2% missing (9 individuals excluded) and (ii) they were related to another individual in the sample (king-cutoff 0.0884) (16 excluded), where one individual from each pair of related samples was excluded based on the King greedy related algorithm. SNPs with high levels of missing data (>3%) (56,268 variants excluded) and a Hardy-Weinberg equilibrium P < 1e-6 were excluded. Samples were further excluded if they were classed as non-European, determined by merging the NCDS combined genotypes with data from 1000 genomes Phase 3), linkage disequilibrium pruning the overlapping single nucleotide polymorphisms (SNPs) such that no pair of SNPs within 50 bp had r2 > 0.20 and visually inspecting the first two genetic principal components along with the known ethnicities of the 1000 genomes sample to define European samples (N=83 excluded).

The final quality controlled imputed set of genotypes contained 6,312 samples and 6,663,631 variants (genome build: GRCh37/ hg19). For comparability with other cohorts, we further restricted to singletons born in Great Britain. 5,989 (37.0%) eligible participants in the 1958c had (valid) genetic data.

### 1970c

The 1970c were genotyped using whole blood samples collected at 46y (Sullivan et al., 2023). Genotyping for 5905 samples (5807 individuals) was performed on the Infinium Global Screening Array-24 v3.0 (consisting of 654,027 genetic variants). One array plate (96 samples) failed during processing and therefore 96 of the samples were repeats. Genotype calling was performed using GenomeStudio (v2.0, Illumina) and quality control was completed using PLINK1.9 and PLINK2.0. 5830 samples were successfully read into GenomeStudio and mapped to a manifest file with the genome build GRCH37. Individuals were excluded if they had (i) they had > 2% missing data (136 samples excluded), (ii) their genotype predicted sex using X chromosome homozygosity was discordant with their reported sex (excluding females with an F value > 0.2 and males with an F value < 0.8) (15 samples excluded), (iii) they had excess heterozygosity [>3 standard deviation (SD) from the mean] (46 samples excluded), (iv) they were related to another individual in the sample (king-cutoff 0.0884) (35 samples excluded), where one individual from each pair of related samples was excluded based on the King greedy related algorithm. The samples which were on the failed array did not pass QC steps and only repeats were kept. The failed samples have been removed from the non QC’d data so that there is only one sample per person. We identified European samples using the GenoPred pipeline which involves (i) merging the BCS70 genotypes with data from 1000 genomes Phase 3, (ii) linkage disequilibrium pruning the overlapping single nucleotide polymorphisms (SNPs) such that no pair of SNPs within 1000 bp had r2 > 0.20 and (iii) using an elastic net model to establish which of the super populations the samples fall into (Africans [AFR], Admixed Americans [AMR], East Asians [EAS], Europeans [EUR] and South Asians [SAS]).

Prior to imputation SNPs with high levels of missing data (>3%), Hardy-Weinberg equilibrium P < 1e-6 or minor allele frequency <1% were excluded. The genetic data were then recoded as vcf files before uploading to the TOPMed Imputation Server which uses Eagle2 to phase haplotypes, and Minimac4 (<https://genome.sph.umich.edu/wiki/Minimac4>) with the TOPMed reference panel. The genome build was updated to hg38 using LiftOver, which is implemented within the TOPMed server. Imputed genotypes were then filtered with PLINK2.0alpha, excluding SNPs with an R2 INFO score < 0.8 and recoded as binary PLINK format. Proceeding with PLINK1.9, samples with >2% missing values, and SNPs with >2 alleles, >3% missing values, Hardy-Weinberg equilibrium P < 1e-6 or a minor allele frequency of <1% were excluded (indels have not been excluded).

The final quality controlled imputed set of genotypes contained 5,598 samples and ~8,640,849 variants and are provided in plink format (genome build: hg38). For comparability with other cohorts, we further restricted to singletons born in Great Britain. 5,170 (31.5%) eligible participants in the 1970c had (valid) genetic data.

### 2001c

The 2001c were genotyped using saliva samples collected at age 14y (Fitzsimons et al., 2022). Genotyping was carried out in the Bristol Genetics Labs (Bristol, UK) using Illumina Infinium global screening arrays-24 v1.0. For more details on the collection of samples, DNA extraction methods, and laboratory procedures, see Fitzsimons et al. (2022). Genotype calling was performed using GenomeStudio (v2.0, Illumina) and quality control was completed using PLINK1.9 and PLINK2.0 (Chang et al., 2015). Samples which could no longer be included in the sample (e.g., due to withdrawn consent) were removed prior to QC. Individuals were excluded if they had > 2% missing data, excess heterozygosity (>3 standard deviation [SD] from the mean), or X chromosome homozygosity discordant with their reported sex (females excluded with an F value > 0.2 and males with an F value < 0.8), so long as these could not be rectified using family relationships inferred using KING.

Prior to imputation, single nucleotide polymorphisms (SNPs) were excluded if they had high levels of missing data (> 3%), Hardy-Weinberg equilibrium P < 1e-6 (based on a subset of unrelated, European samples), or minor allele frequency (MAF) < 1%. The genetic data were then recoded as .vcf files before uploading to the TOPMed Imputation Server, which uses Eagle2 to phase haplotypes and Minimac4 (<https://genome.sph.umich.edu/wiki/Minimac4>) with the TOPMed reference panel. Imputed genotypes were then filtered with PLINK2.0alpha, excluding SNPs with an R2 INFO score < 0.8, and recoded as hard-calls into binary PLINK format. Proceeding with PLINK1.9, samples with > 2% missing values were removed and SNPs were excluded if they had > 3% missing values, > 2 alleles or a MAF of < 1%. Duplicate samples were also removed, with the sample with the higher genotyping rate retained. In these steps, European individuals were identified using the GenoPred pipeline which involved (a) merging the MCS genotypes with data from 1000 genomes Phase 3, (b) linkage disequilibrium pruning overlapping SNPs such that no pair of SNPs within 1000 bp had r^2^ > 0.20 and (c) using an elastic net model to identify Europeans versus non-Europeans.

For comparability with other cohorts, we further restricted to singletons born in Great Britain. 5,489 (41.1%) eligible participants in the 2001c had (valid) genetic data.

#### Measures

##### Covariates

We included a range of variables as covariates in regression models, predictors of selection into the genotyped sample, and auxiliary variables in multiple imputation models. These were cohort member’s sex, verbal cognitive ability at age 10/11, maternal age at birth, mother’s and father’s years of education, family socioeconomic class, mother’s and father’s BMI, and cohort member’s first ten genetic principal components (PCs).

Verbal cognitive ability was measured at age 11 in the 1946c and 1958c using the verbal items from the General Ability Test (GAT). Verbal cognitive ability was measured in the 1970c and 2001c at ages 10 and 11, respectively, using the Word Similarities test from British Ability Scales (BAS). For comparability we standardised this variable in each cohort (mean = 0, SD = 1).

Mother’s and father’s years of education were calculated using age at leaving education. We used data from natural parent’s where information on parent type was available. In some instances, information on parental education was collected in multiple survey sweeps. Where this was the case, we used data from the earliest possible sweep.

Family socioeconomic class was measured using the Registrar General Social Class (RGSC) schema in the 1946c, 1958c and 1970c and using the National Statistics Socio-Economic Classification (NS-SEC) schema in the 2001c. For the 1946c, we used father’s social class when cohort members were age 4y, or if missing, 11y. For the 1958c, we used social class of the male household head collected when cohort members were age 7y or, if missing, 11y. For the 1970c, we used the higher of mother’s and father’s social class when cohort members were age 10y. For the 2001c, social class we used highest parental or guardian social class. As this data was available in multiple sweeps, used the earliest sweep for which data were available.

Mother’s and father’s BMI were measured differently in each cohort. In each case, only self-reported height and weight were available. Data collection was at ages 6y, 11y, and 10y for the 1946c, 1958c and 1970c, respectively. Though, for the 1958c and 1970c, mother’s height measurements were also available from the cohort member’s birth; we used these if later height data was missing. For the 2001c, questions on height and weight were asked at multiple sweeps. We used the earliest available measurement per parent.

#### Statistical Analysis

##### Derivation of Inverse Probability Weights for Selection into the Genotyped Sample

Genotyped samples were taken in the 1946c, 1958c, 1970c and 2001c at ages 53y, 44y, and 46y and 14y, respectively. There was attrition in each cohort over time and not all participants asked agreed to give a DNA sample or gave a sample that passed QC. Selection into the genotyped sample was not random (Table S2). To try to account for this, we created inverse probability weights for each cohort’s genotyped sample so that, reweighted, they had similar (observed) characteristics to the larger set of recruited participants.

To create the weights, for each cohort, we regressed selection into the genotyped sample on a set of predictor variables using logistic regression. We used these models to obtain a predicted probability of selection into the genotyped sample and then for each individual in the genotyped sample took the inverse of this probability to create a response weight, scaling the weights so their sum equalled the size of the genotyped sample. As the 1946c and 2001c each had design weights to account for unequal selection into the baseline sample, we combined design and genetic-response weights by multiplying them together prior to scaling.

The variables we used to predict participation into the genotyped sample were sex, family socioeconomic class, mother’s and father’s year of education, mother’s and father’s BMI, verbal cognitive ability @ age 10/11, and mother’s and father’s age at participant’s birth. (Father’s age at birth was not available in the 1946c.) Each of these variables was collected during the participant’s childhood or adolescence. We chose early measures to reduce missingness in the predictor variables. Nevertheless, some missingness remained so we imputed missing values using multiple imputation (Classification and Regression Trees; 50 imputations). Weights were calculated in each imputed dataset and then averaged to create a final weight that we used in subsequent substantive analyses.

There was also missingness in the data among the genotyped samples used in substantive analysis – there was missingness in covariates as well as a recorded BMI. After creating weights for the genotyped sample, we used multiple imputation within this sample to impute remaining missing values (Classification and Regression Trees; 50 imputations). In sensitivity analyses which used the weighted and imputed data, we pooled results using Rubin’s Rules (Rubin, 1987).

### Results

#### Tables

Table S1: Association between childhood PGI and covariates

|  | Variable | 1946c | 1958c | 1970c | 2001c |
| --- | --- | --- | --- | --- | --- |
| Sex | Male | -0.02 (0.98) | 0.00 (1.01) | 0.04 (1.00) | -0.03 (1.00) |
|  | Female | 0.02 (1.03) | -0.02 (1.00) | -0.01 (0.99) | 0.02 (0.99) |
|  | Verbal Score @ Age 10/11 | -0.01 (-0.06, 0.04) | -0.01 (-0.04, 0.02) | 0.02 (-0.01, 0.05) | 0.00 (-0.03, 0.03) |
|  | Mother's BMI | 0.12* (0.08, 0.17) | 0.08* (0.05, 0.11) | 0.08* (0.05, 0.11) | 0.09* (0.05, 0.12) |
|  | Father's BMI | 0.05 (-0.01, 0.10) | 0.06* (0.03, 0.09) | 0.06* (0.03, 0.10) | 0.10* (0.06, 0.13) |
|  | Mother's Age | -0.02 (-0.07, 0.03) | -0.01 (-0.04, 0.02) | -0.02 (-0.04, 0.01) | 0.02 (-0.01, 0.06) |
|  | Mother's Education (Years) | 0.02 (-0.02, 0.06) | 0.02 (-0.01, 0.05) | -0.01 (-0.04, 0.02) | -0.01 (-0.04, 0.02) |
|  | Father's Education (Years) | 0.01 (-0.03, 0.05) | 0.01 (-0.02, 0.04) | -0.01 (-0.04, 0.02) | -0.01 (-0.04, 0.02) |
| Family Class (Registrar General) | I Professional | -0.10 (0.99) | -0.01 (1.02) | 0.01 (0.95) | -0.02 (0.97) |
|  | II Intermediate | 0.13 (1.02) | -0.03 (1.01) | -0.02 (0.97) | -0.02 (0.98) |
|  | III Skilled Manual | -0.01 (1.05) | -0.01 (0.96) | -0.01 (0.99) | 0.00 (1.01) |
|  | III Skilled Non-Manual | 0.02 (1.01) | -0.03 (1.01) | 0.05 (1.02) | 0.16 (0.99) |
|  | IV Semi-Skilled | -0.05 (0.96) | 0.01 (1.02) | 0.02 (1.04) | 0.04 (1.02) |
|  | V Unskilled | -0.10 (0.98) | 0.09 (0.97) | -0.18 (1.09) | -0.04 (0.97) |
|  | Not Working |  |  |  | -0.08 (1.00) |
| Mean (SD) for categorical covariates, and Pearson’s correlation (+ 95% CIs) for continuous covariates.  * indicates variable is a statistically significant predictor of the PGI (Wald test, p < 0.05).  Estimates were weighted using recruitment weights and account for complex survey design. Row-wise complete case data.  Full definitions for each variable are provided above. | | | | | |

Table S2: Descriptive statistics according to whether the participant was genotyped or was part of eligible sample (singleton of White ethnicity, born in England, Scotland or Wales). Row-wise complete case data, weighted accounting for complex survey design.

|  | | 1946c | | 1958c | | 1970c | | 2001c | |
| --- | --- | --- | --- | --- | --- | --- | --- | --- | --- |
|  | Variable | Eligible | Genotyped | Eligible | Genotyped | Eligible | Genotyped | Eligible | Genotyped |
| Sex | Male | 2,818.8 (52.6%) | 1,374.7 (50.3%)* | 8,330.0 (51.5%) | 3,000.0 (50.1%)* | 8,512.0 (51.9%) | 2,536.0 (49.1%)* | 6,857.0 (51.3%) | 2,741.4 (50.0%)* |
|  | Female | 2,542.3 (47.4%) | 1,356.3 (49.7%) | 7,844.0 (48.5%) | 2,989.0 (49.9%) | 7,890.0 (48.1%) | 2,634.0 (50.9%) | 6,509.2 (48.7%) | 2,746.8 (50.0%) |
|  | Verbal Score @ Age 10/11 | 0.0 (1.0) | 0.1 (1.0)* | 0.0 (1.0) | 0.1 (1.0)* | 0.0 (1.0) | 0.2 (0.9)* | 0.0 (1.0) | 0.1 (0.9)* |
|  | Mother's BMI | 23.4 (3.8) | 23.5 (3.8) | 23.7 (3.7) | 23.6 (3.7)* | 23.2 (3.3) | 23.0 (3.2)* | 24.7 (4.4) | 24.8 (4.5) |
|  | Father's BMI | 23.9 (2.9) | 23.9 (2.9) | 24.7 (3.0) | 24.6 (2.9) | 24.5 (2.8) | 24.3 (2.7)* | 26.0 (3.5) | 26.1 (3.5)* |
|  | Mother's Age | 28.8 (5.8) | 28.7 (5.8) | 27.4 (5.7) | 27.4 (5.6) | 25.9 (5.5) | 25.9 (5.2) | 29.4 (5.9) | 30.0 (5.6)* |
|  | Mother's Education (Years) | 3.4 (1.0) | 3.4 (1.0) | 3.9 (1.3) | 4.0 (1.4)* | 4.6 (1.4) | 4.8 (1.4)* | 6.2 (1.7) | 6.5 (1.8)* |
|  | Father's Education (Years) | 3.4 (1.1) | 3.4 (1.1) | 3.9 (1.5) | 4.0 (1.6)* | 4.8 (1.7) | 4.9 (1.7)* | 6.1 (1.8) | 6.2 (1.8)* |
| Family Class (Registrar General) | I Professional | 128.1 (2.7%) | 74.2 (2.9%) | 758.0 (5.2%) | 344.0 (6.0%)* | 721.0 (6.0%) | 328.0 (7.3%)* | 637.1 (6.8%) | 409.8 (7.8%)* |
|  | II Intermediate | 549.6 (11.7%) | 337.2 (13.0%) | 2,151.0 (14.8%) | 911.0 (15.9%) | 3,438.0 (28.4%) | 1,433.0 (32.0%) | 4,265.8 (45.5%) | 2,523.5 (48.3%) |
|  | III Skilled Non-Manual | 470.4 (10.0%) | 273.5 (10.5%) | 1,444.0 (10.0%) | 568.0 (9.9%) | 2,571.0 (21.2%) | 955.0 (21.3%) | 1,392.7 (14.9%) | 743.0 (14.2%) |
|  | III Skilled Manual | 2,137.7 (45.6%) | 1,150.4 (44.2%) | 6,569.0 (45.3%) | 2,603.0 (45.6%) | 3,759.0 (31.1%) | 1,268.0 (28.3%) | 945.7 (10.1%) | 522.1 (10.0%) |
|  | IV Semi-Skilled | 946.1 (20.2%) | 527.3 (20.3%) | 2,593.0 (17.9%) | 973.0 (17.0%) | 1,302.0 (10.8%) | 407.0 (9.1%) | 770.6 (8.2%) | 406.8 (7.8%) |
|  | V Unskilled | 451.1 (9.6%) | 238.4 (9.2%) | 976.0 (6.7%) | 315.0 (5.5%) | 311.0 (2.6%) | 86.0 (1.9%) | 118.8 (1.3%) | 60.5 (1.2%) |
|  | Not Working |  |  |  |  |  |  | 1,246.4 (13.3%) | 557.6 (10.7%) |

Table S3: Heritability of BMI and association between adulthood and childhood PGI and BMI by cohort and follow-up.

|  | | Association Between PGI & BMI | | PGI-Heritability (Incremental R^2^) | |  |
| --- | --- | --- | --- | --- | --- | --- |
| Cohort | Age | (a) Adulthood PGI | (b) Childhood PGI | (c) Adulthood PGI | (d) Childhood PGI | (e) SNP-Heritability |
| 1946c | 4 | 0.14 (0.06, 0.23) | 0.22 (0.14, 0.29) | 0.9% (0.1%, 2.1%) | 2.0% (0.8%, 3.5%) | 21.0% (-7.8%, 49.8%) |
|  | 6 | 0.14 (0.07, 0.20) | 0.19 (0.13, 0.26) | 1.2% (0.2%, 2.3%) | 2.3% (1.0%, 4.0%) | 35.7% (5.1%, 66.2%) |
|  | 7 | 0.14 (0.08, 0.21) | 0.21 (0.14, 0.27) | 1.3% (0.4%, 2.5%) | 2.5% (1.2%, 4.1%) | 15.4% (-14.5%, 45.2%) |
|  | 11 | 0.37 (0.25, 0.48) | 0.51 (0.39, 0.62) | 2.6% (1.4%, 4.2%) | 5.1% (3.1%, 7.2%) | 42.3% (12.5%, 72.1%) |
|  | 15 | 0.52 (0.38, 0.65) | 0.55 (0.43, 0.68) | 4.0% (2.2%, 6.1%) | 4.6% (2.7%, 6.8%) | 45.5% (13.9%, 77.2%) |
|  | 26 | 0.50 (0.37, 0.63) | 0.45 (0.33, 0.58) | 3.2% (1.8%, 5.0%) | 2.7% (1.4%, 4.2%) | 30.0% (0.7%, 59.3%) |
|  | 36 | 0.63 (0.49, 0.76) | 0.48 (0.35, 0.61) | 4.0% (2.4%, 5.7%) | 2.4% (1.2%, 3.8%) | 49.5% (21.5%, 77.4%) |
|  | 43 | 0.72 (0.55, 0.88) | 0.49 (0.33, 0.66) | 4.1% (2.5%, 5.9%) | 2.0% (0.9%, 3.2%) | 45.8% (19.1%, 72.6%) |
|  | 53 | 0.80 (0.60, 0.99) | 0.53 (0.34, 0.73) | 3.5% (2.2%, 5.2%) | 1.6% (0.7%, 2.8%) | 14.5% (-11.0%, 40.0%) |
|  | 63 | 0.78 (0.55, 1.01) | 0.22 (0.00, 0.45) | 3.1% (1.5%, 5.0%) | 0.3% (0.0%, 1.0%) | 39.5% (5.9%, 73.0%) |
|  | 69 | 0.73 (0.48, 0.98) | 0.23 (-0.02, 0.48) | 2.6% (1.1%, 4.4%) | 0.3% (0.0%, 1.1%) | 47.4% (12.3%, 82.5%) |
| 1958c | 7 | 0.17 (0.12, 0.21) | 0.28 (0.23, 0.32) | 1.1% (0.6%, 1.6%) | 3.0% (2.2%, 3.9%) | 17.8% (4.0%, 31.6%) |
|  | 11 | 0.36 (0.29, 0.43) | 0.47 (0.40, 0.54) | 2.2% (1.4%, 3.0%) | 3.7% (2.7%, 4.8%) | 35.1% (20.5%, 49.7%) |
|  | 16 | 0.38 (0.30, 0.46) | 0.51 (0.43, 0.58) | 2.1% (1.3%, 2.9%) | 3.8% (2.8%, 4.9%) | 22.9% (7.0%, 38.7%) |
|  | 23 | 0.48 (0.41, 0.56) | 0.50 (0.43, 0.57) | 3.1% (2.3%, 4.0%) | 3.4% (2.5%, 4.4%) | 27.4% (13.7%, 41.0%) |
|  | 33 | 0.64 (0.54, 0.74) | 0.57 (0.47, 0.67) | 2.9% (2.0%, 3.8%) | 2.4% (1.6%, 3.2%) | 21.6% (8.4%, 34.8%) |
|  | 42 | 0.74 (0.64, 0.84) | 0.53 (0.43, 0.63) | 3.5% (2.7%, 4.4%) | 1.9% (1.3%, 2.5%) | 19.7% (7.5%, 32.0%) |
|  | 44 | 0.87 (0.75, 0.98) | 0.59 (0.47, 0.71) | 3.6% (2.8%, 4.6%) | 1.7% (1.1%, 2.4%) | 19.7% (8.1%, 31.3%) |
|  | 50 | 0.88 (0.76, 1.01) | 0.52 (0.40, 0.65) | 3.7% (2.7%, 4.6%) | 1.3% (0.7%, 2.0%) | 14.5% (1.1%, 27.9%) |
|  | 55 | 0.83 (0.70, 0.96) | 0.42 (0.28, 0.55) | 3.2% (2.2%, 4.2%) | 0.8% (0.4%, 1.4%) | 23.6% (8.5%, 38.6%) |
| 1970c | 10 | 0.31 (0.25, 0.37) | 0.45 (0.39, 0.51) | 2.4% (1.6%, 3.3%) | 5.0% (3.7%, 6.3%) | 27.7% (11.1%, 44.2%) |
|  | 16 | 0.52 (0.41, 0.62) | 0.54 (0.45, 0.64) | 3.2% (2.0%, 4.6%) | 3.7% (2.6%, 5.0%) | 20.1% (-2.2%, 42.4%) |
|  | 29 | 0.76 (0.65, 0.86) | 0.64 (0.53, 0.74) | 3.9% (2.9%, 5.0%) | 2.8% (2.0%, 3.8%) | 27.5% (11.6%, 43.3%) |
|  | 34 | 0.80 (0.68, 0.92) | 0.67 (0.55, 0.79) | 3.8% (2.8%, 4.9%) | 2.7% (1.9%, 3.7%) | 19.3% (3.0%, 35.6%) |
|  | 42 | 0.84 (0.72, 0.97) | 0.64 (0.52, 0.77) | 3.4% (2.5%, 4.4%) | 2.1% (1.4%, 2.9%) | 21.7% (6.4%, 37.0%) |
|  | 46 | 0.99 (0.85, 1.13) | 0.67 (0.53, 0.80) | 3.7% (2.8%, 4.7%) | 1.7% (1.1%, 2.4%) | 21.8% (8.3%, 35.4%) |
| 2001c | 3 | 0.09 (0.05, 0.13) | 0.22 (0.17, 0.27) | 0.5% (0.2%, 0.9%) | 2.7% (1.7%, 3.8%) | 30.7% (16.3%, 45.2%) |
|  | 5 | 0.17 (0.13, 0.21) | 0.29 (0.25, 0.34) | 1.4% (0.8%, 2.1%) | 4.1% (2.9%, 5.4%) | 28.6% (15.3%, 41.9%) |
|  | 7 | 0.32 (0.27, 0.37) | 0.47 (0.41, 0.54) | 2.5% (1.7%, 3.3%) | 5.3% (4.0%, 6.8%) | 32.8% (19.0%, 46.5%) |
|  | 11 | 0.63 (0.54, 0.73) | 0.76 (0.65, 0.88) | 3.6% (2.6%, 4.6%) | 5.0% (3.7%, 6.2%) | 34.9% (21.1%, 48.7%) |
|  | 14 | 0.74 (0.64, 0.85) | 0.85 (0.73, 0.97) | 3.8% (2.8%, 4.9%) | 4.7% (3.4%, 5.9%) | 42.7% (29.3%, 56.1%) |
|  | 17 | 0.93 (0.80, 1.06) | 1.04 (0.88, 1.20) | 4.3% (3.2%, 5.5%) | 5.2% (3.6%, 6.9%) | 34.2% (17.9%, 50.5%) |
| Columns A-B: Regression of BMI (kg/m^2^) upon PGI by cohort, age of follow-up, and PGI (adulthood or childhood). Derived from separate OLS regressions adjusting for age, sex and first 10 genetic principal components. Estimates were weighted using recruitment weights and account for complex survey design. Marginal effect of 1 SD higher PGI and BMI (kg/m^2^).  Columns C-D: incremental proportion of variance explained by (adulthood or childhood) PGI calculated comparing with regression of BMI on age, sex and first 10 genetic principal components. Confidence intervals estimated using bootstrapping (500 bootstraps, percentile method).  Column E: SNP-heritability of BMI calculated with GCTA, adjusting for sex, age and first 10 genetic principal components. Survey weights were not incorporated in this analysis. | | | | | | |

#### Figures

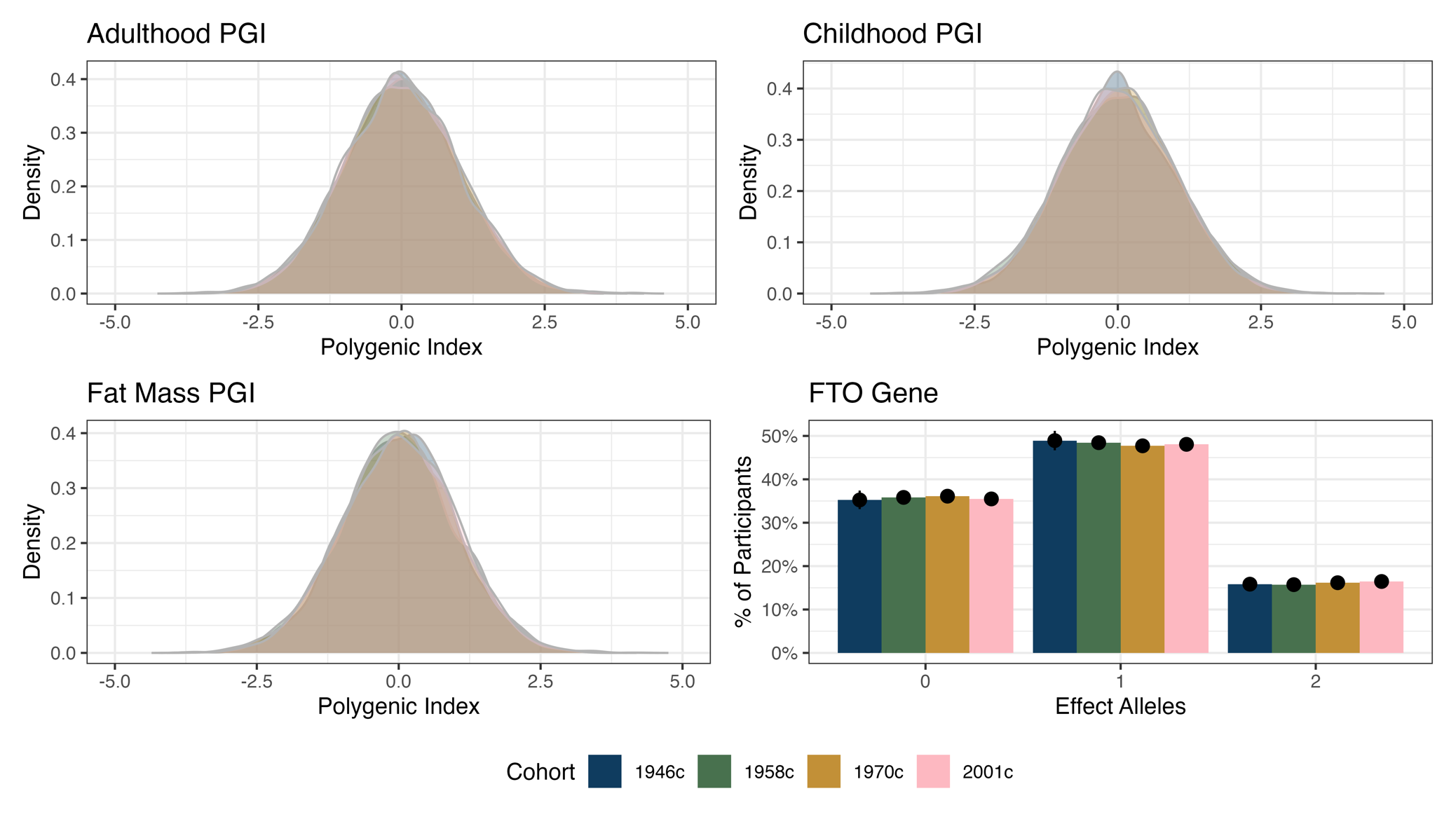

Figure S1: Distribution of PGI and rs1558902 FTO variant by cohort. Weighted using recruitment weights.

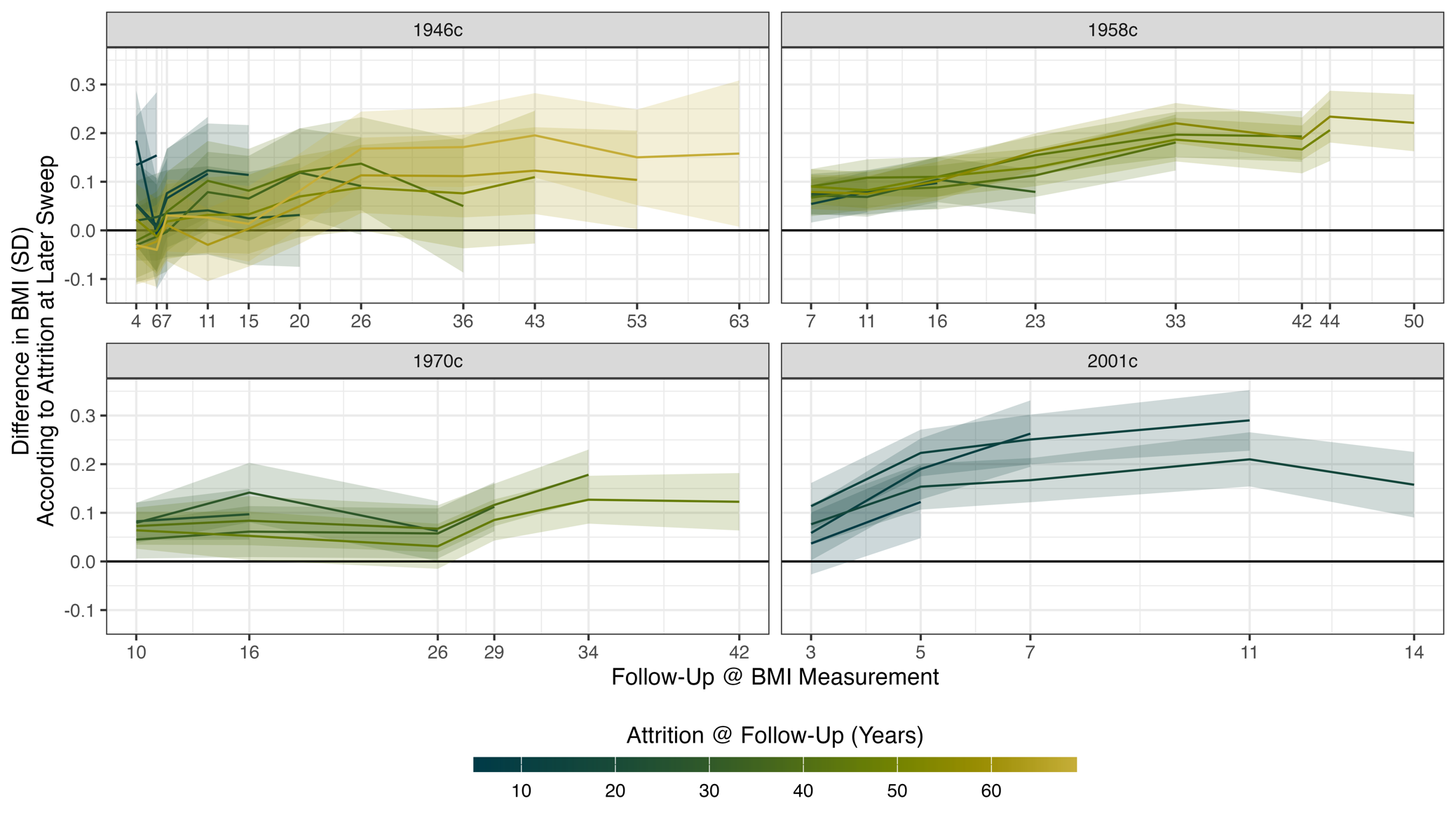

Figure S2: Difference in BMI (SD) according to drop-out at a later sweep by cohort, age of BMI measurement, and age at which drop-out was assessed. X-axis displays the age at which BMI was measured. Line color indicates the age at which drop-out was determined. Unweighted data.

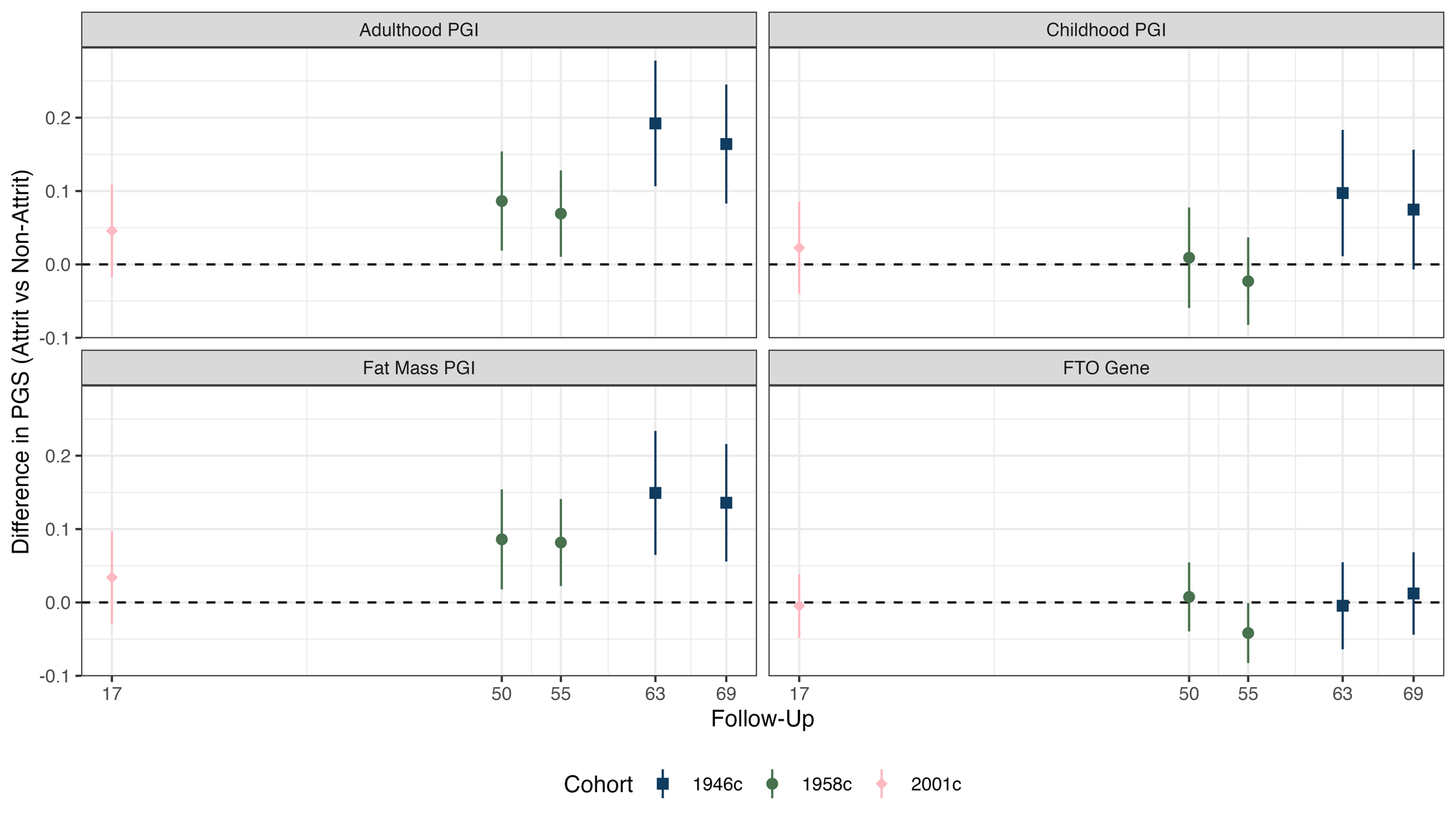

Figure S3: Difference in PGI according to drop-out at a sweep following the collection of DNA by cohort, PGI, and age at which drop-out was assessed. X-axis displays the age at which drop-out was determined. Unweighted data. Collection of DNA was at age 53y in the 1946c, 42y in the 1958c, 46y in the 1970c and 14y in the 2001c.

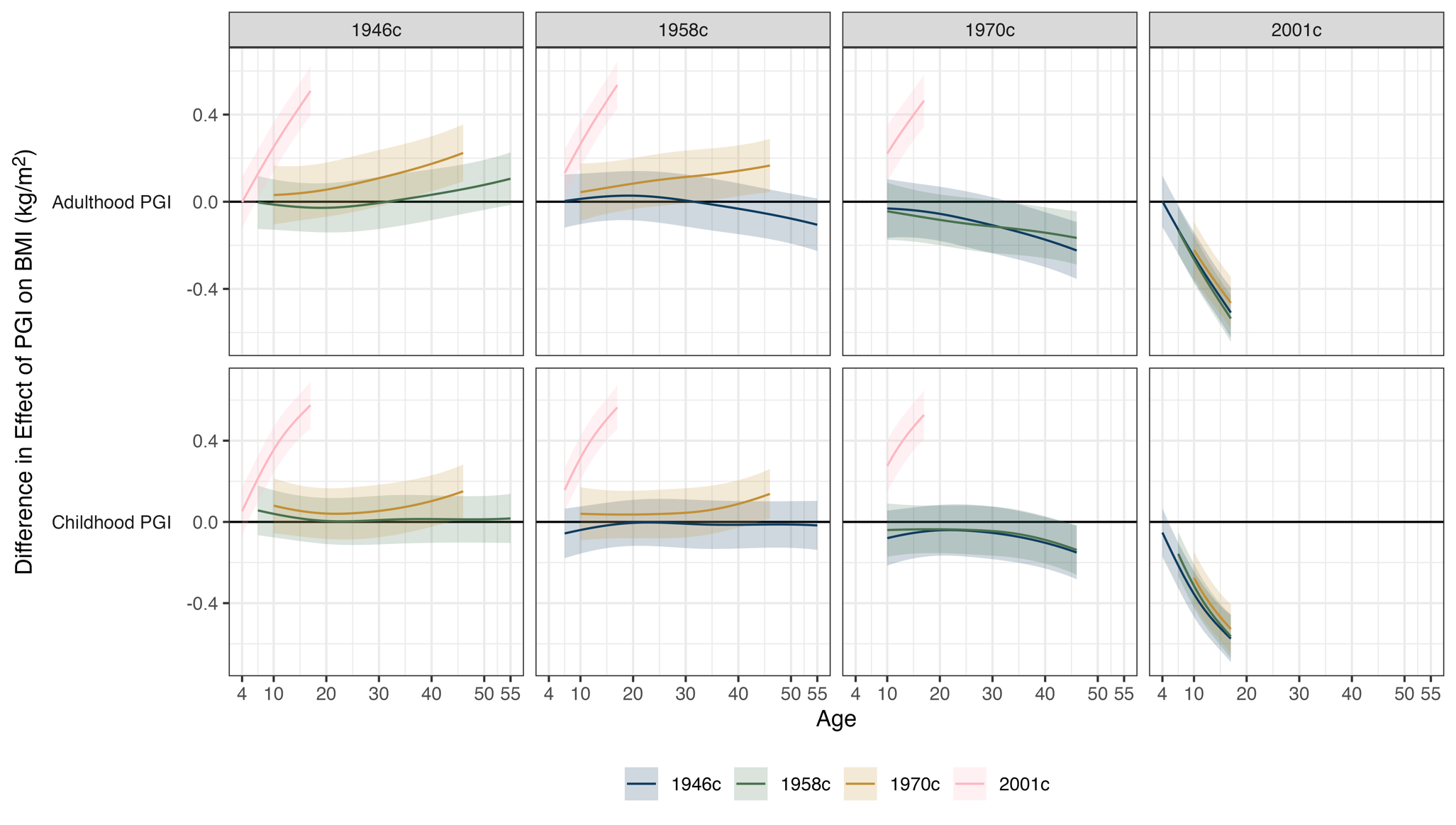

Figure S4: Difference in association between PGI and BMI (kg/m^2^) by age and PGI (adulthood or childhood) for specified pair of cohorts. Derived by estimating marginal effects from cohort-specific linear mixed effects models and then calculating z-scores for differences in these marginal effects. In linear mixed effect models, the association between PGI and BMI allowed to vary by age (two natural splines) with adjustment for age (two natural splines), sex, first 10 genetic principal components, and person-specific random intercept. Difference shown is marginal effect for cohort represented by line minus cohort list in plot header. For instance, a positive value indicates the association between PGI and BMI was stronger in the line-cohort than the plot-header cohort. Estimates were weighted using recruitment weights.

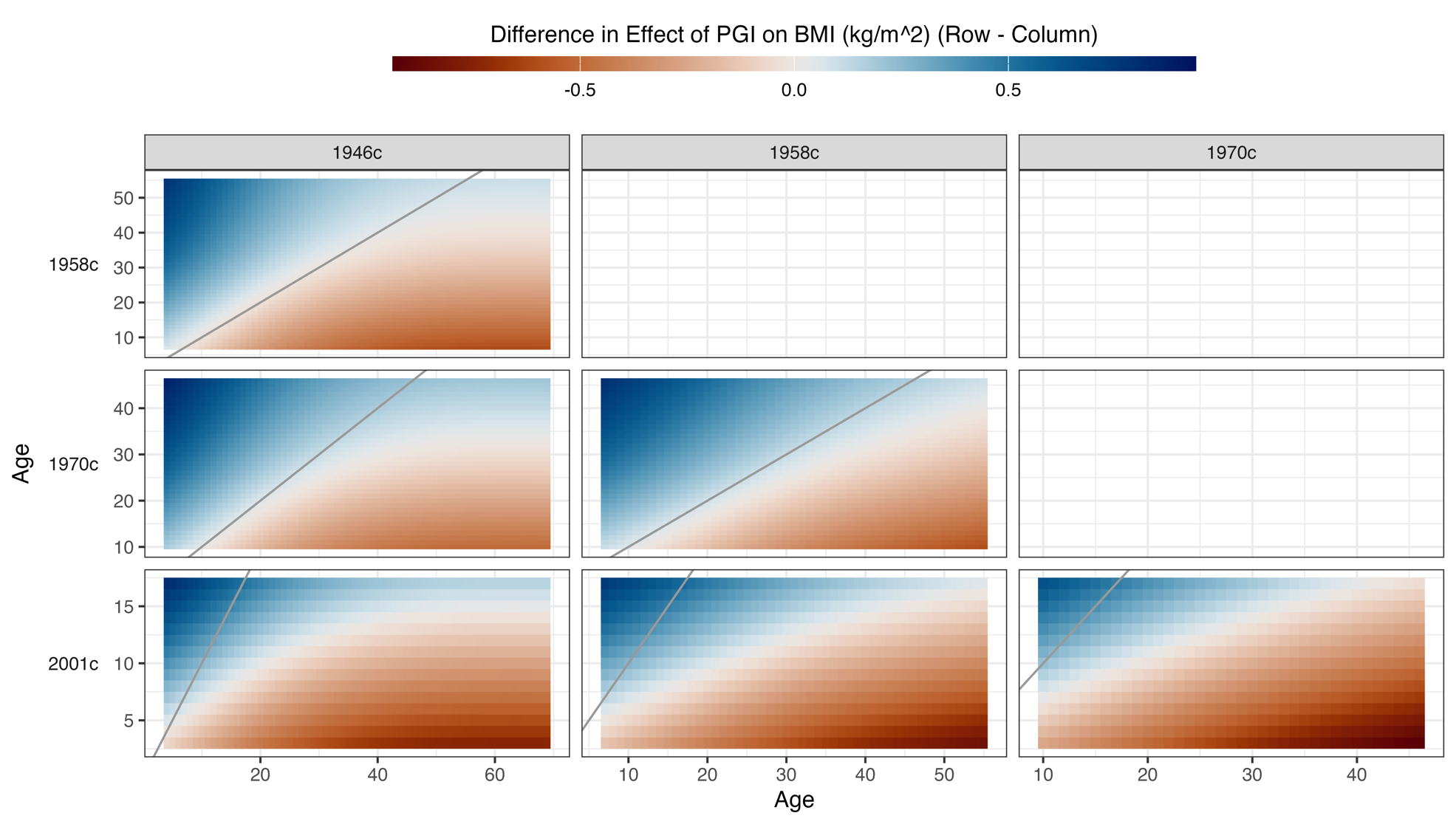

Figure S5: Contour plot of difference in association between adulthood PGI and BMI (kg/m^2^) for specified pair of cohorts at specified combination of ages. Derived by estimating marginal effects from cohort-specific linear mixed effects models and then calculating z-scores for differences in these marginal effects. In linear mixed effect models, the association between PGI and BMI allowed to vary by age (two natural splines) with adjustment for age (two natural splines), sex, first 10 genetic principal components, and person-specific random intercept. Difference shown is marginal effect for the cohort listed in the row minus cohort list in plot header. For instance, a positive value (blue shading) indicates the association between PGI and BMI was stronger in the row-cohort than the plot-header cohort. Absolute line (grey) added for reference – blue values to the right of this line imply row-cohort has stronger association at earlier age than plot-header cohort. Estimates were weighted using recruitment weights.

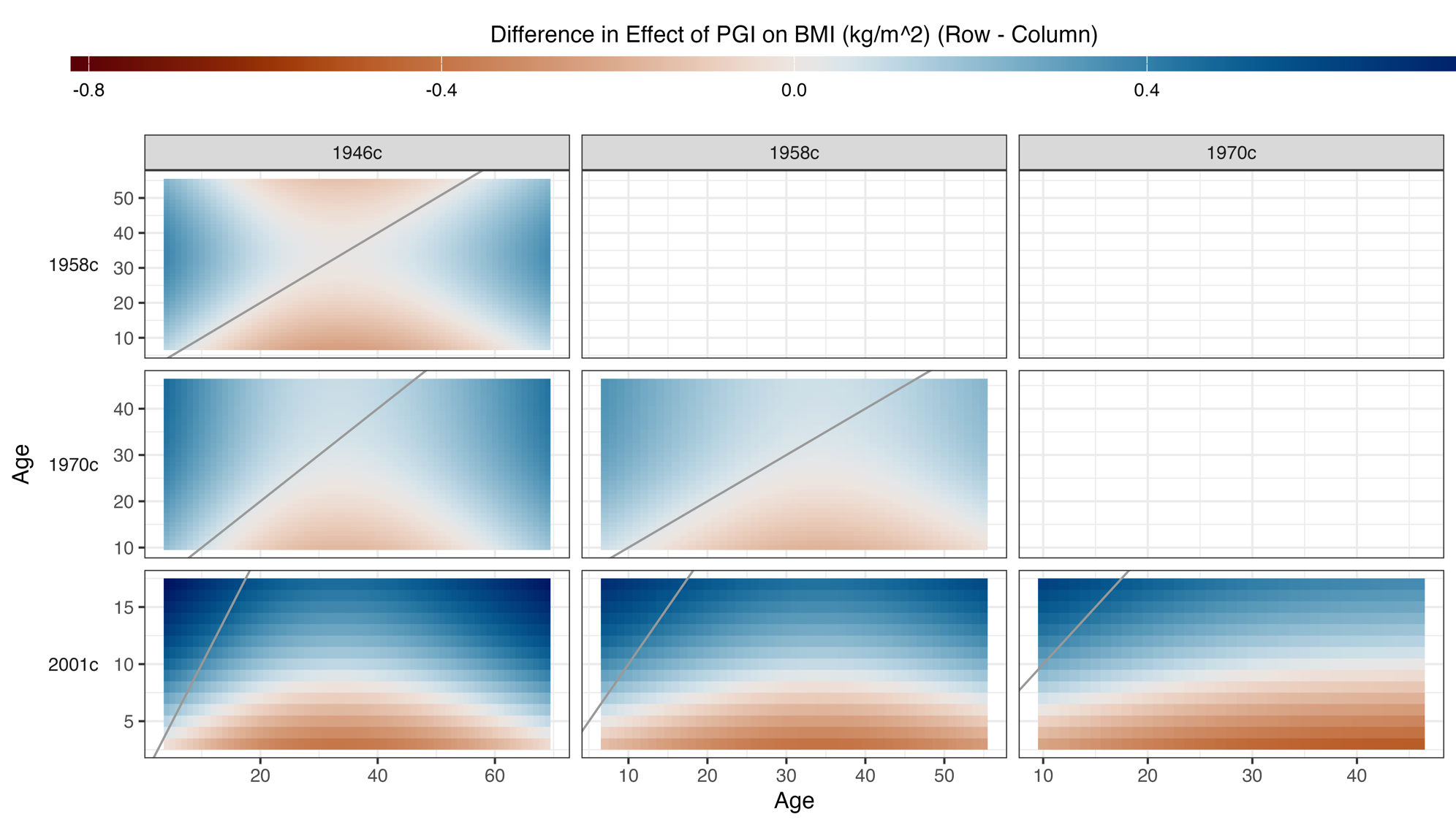

Figure S6: Contour plot of difference in association between childhood PGI and BMI (kg/m^2^) for specified pair of cohorts at specified combination of ages. Derived by estimating marginal effects from cohort-specific linear mixed effects models and then calculating z-scores for differences in these marginal effects. In linear mixed effect models, the association between PGI and BMI allowed to vary by age (two natural splines) with adjustment for age (two natural splines), sex, first 10 genetic principal components, and person-specific random intercept. Difference shown is marginal effect for the cohort listed in the row minus cohort list in plot header. For instance, a positive value (blue shading) indicates the association between PGI and BMI was stronger in the row-cohort than the plot-header cohort. Absolute line (grey) added for reference – blue values to the right of this line imply row-cohort has stronger association at earlier age than plot-header cohort. Estimates were weighted using recruitment weights.

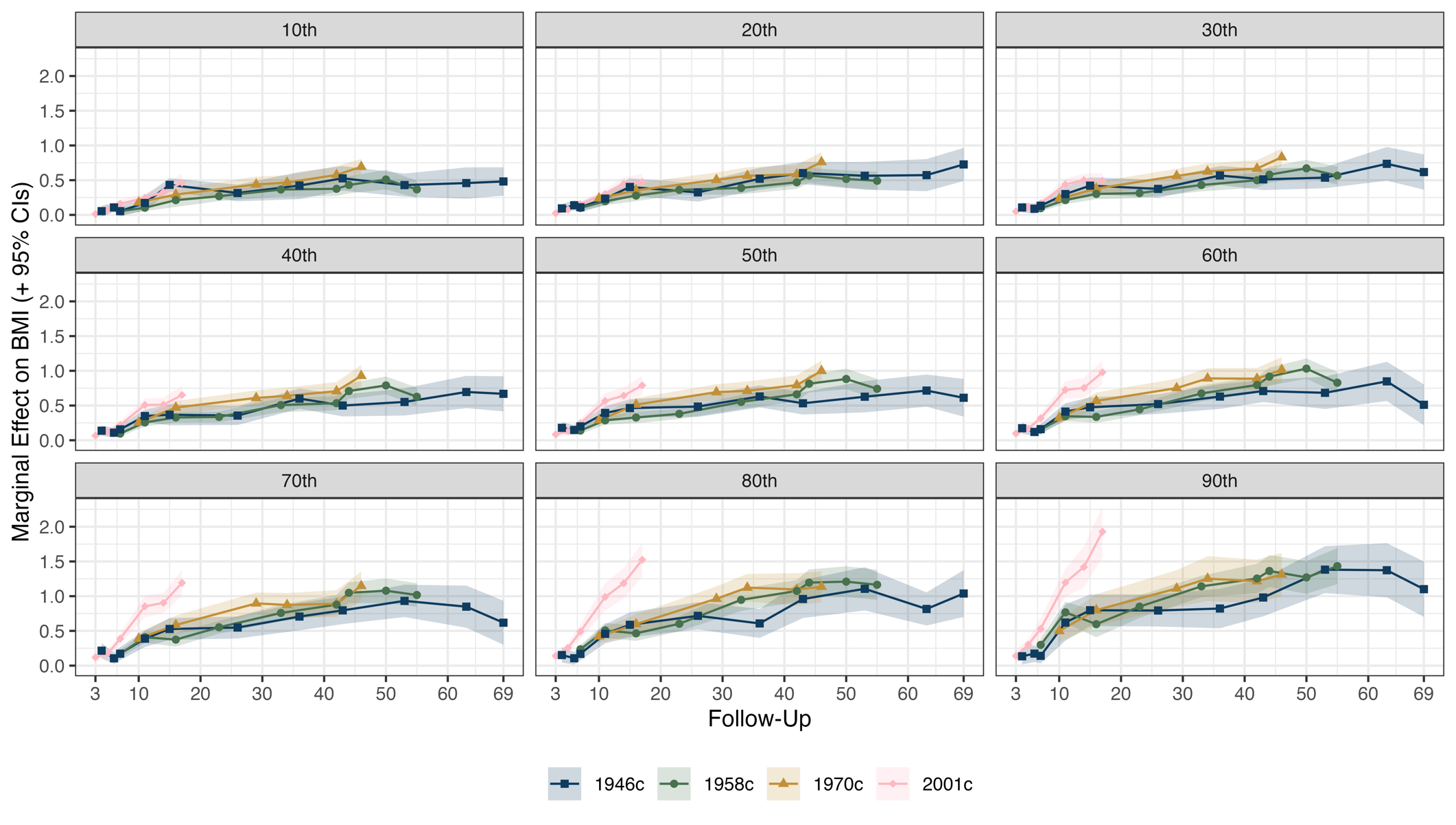

Figure S7: Association between adulthood PGI and BMI (kg/m^2^) by BMI decile, cohort, age of follow-up. Derived from separate quantile regressions adjusting for age (linear term), sex and first 10 genetic principal components. Estimates were weighted using recruitment weights. Each panels displays associations for a particular decile of BMI (10^th^, 20^th^, …, 90^th^). Results show how the (conditional) centiles of BMI vary according to 1 SD increases in the adulthood PGI.

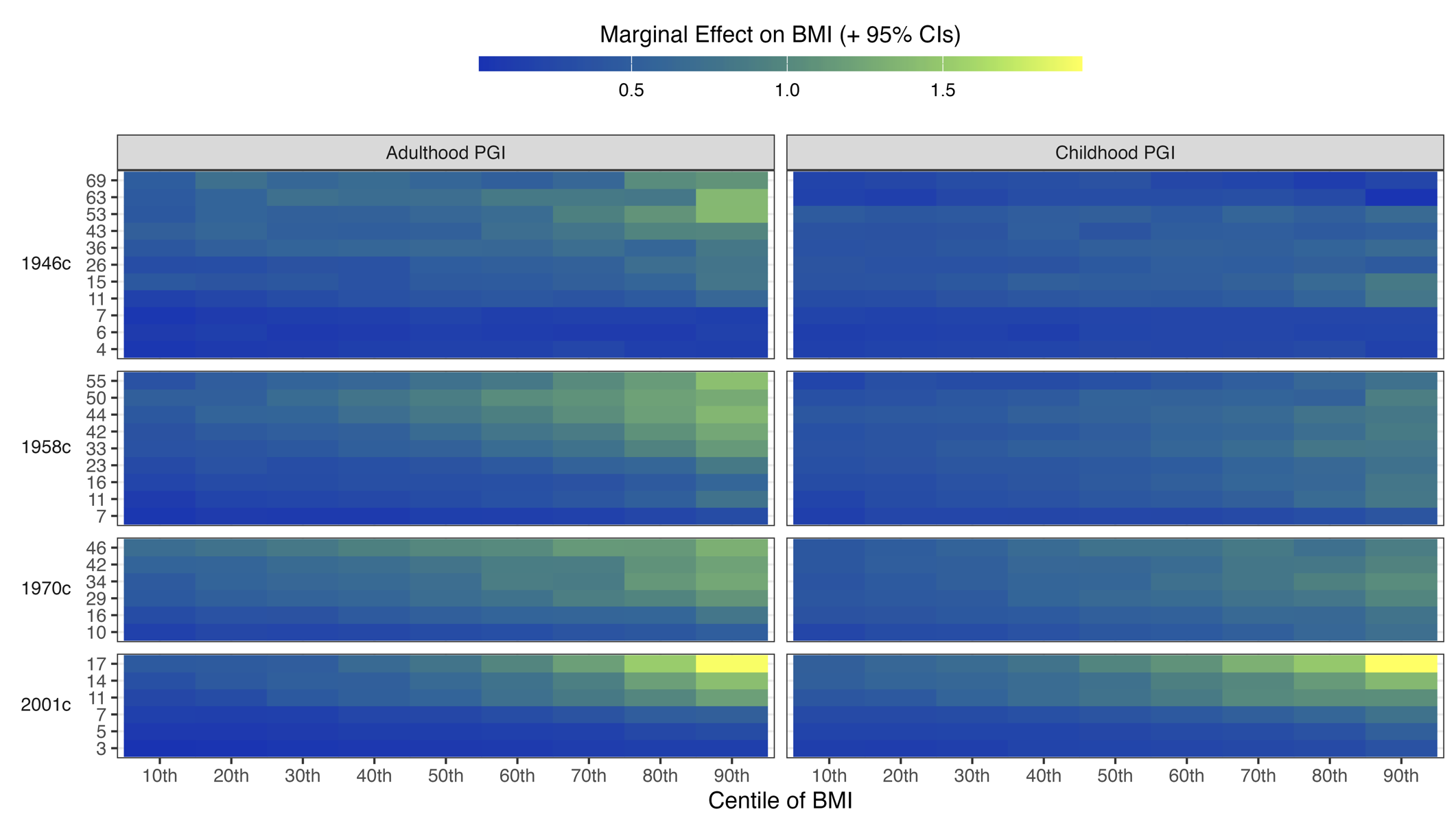

Figure S8: Heatmaps of association between adulthood PGI and BMI (kg/m^2^) by BMI decile, cohort, age of follow-up. Derived from separate quantile regressions adjusting for age (linear term), sex and first 10 genetic principal components. Estimates were weighted using recruitment weights. Each panels displays associations for a particular decile of BMI (10^th^, 20^th^, …, 90^th^) with the size of the association represented by the shading of the cell. Results show how the (conditional) centiles of BMI vary according to 1 SD increases in the adulthood PGI.

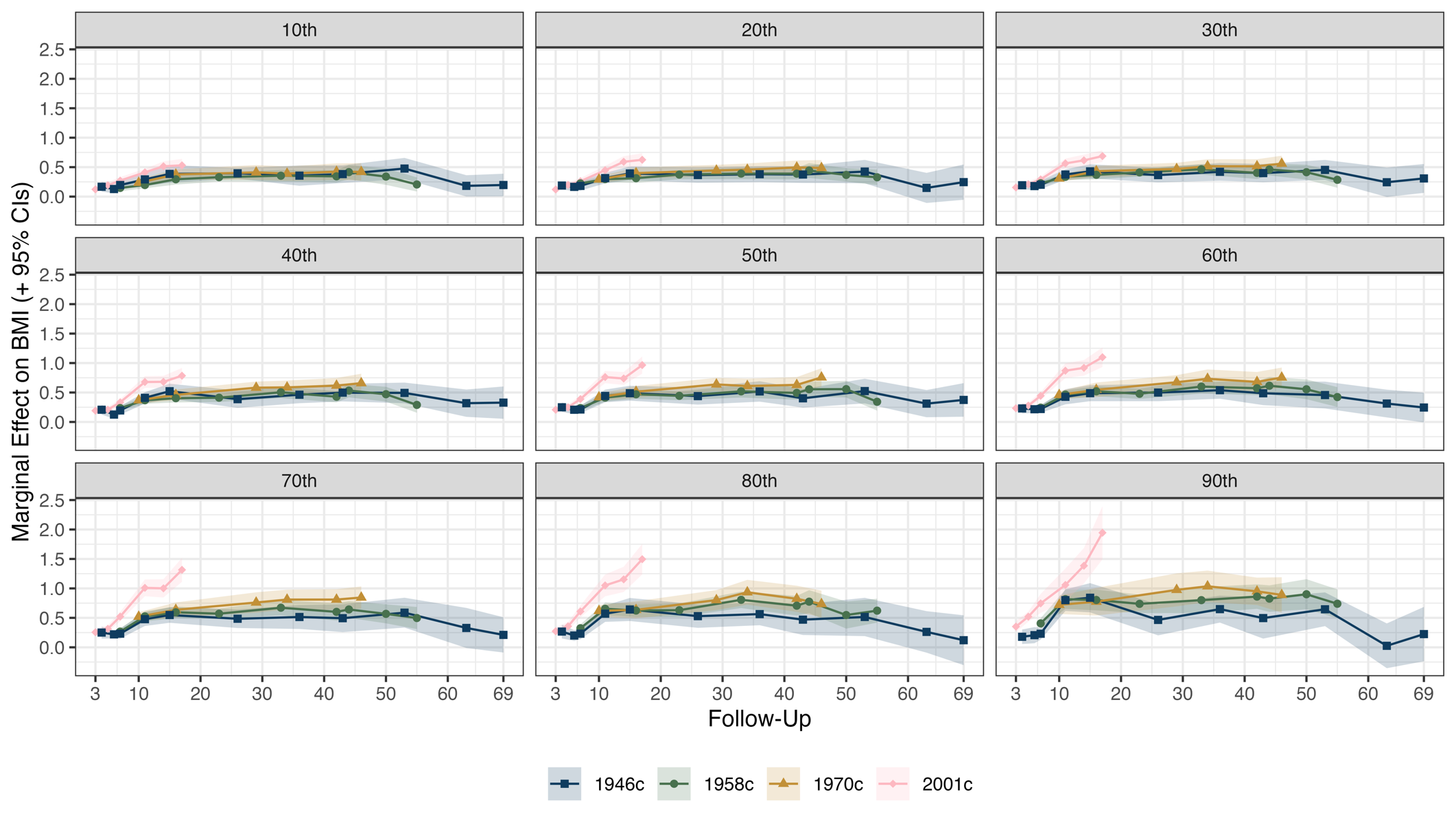

Figure S9: Association between childhood PGI and BMI (kg/m^2^) by BMI decile, cohort, age of follow-up. Derived from separate quantile regressions adjusting for age (linear term), sex and first 10 genetic principal components. Estimates were weighted using recruitment weights. Each panels displays associations for a particular decile of BMI (10^th^, 20^th^, …, 90^th^). Results show how the (conditional) centiles of BMI vary according to 1 SD increases in the adulthood PGI.

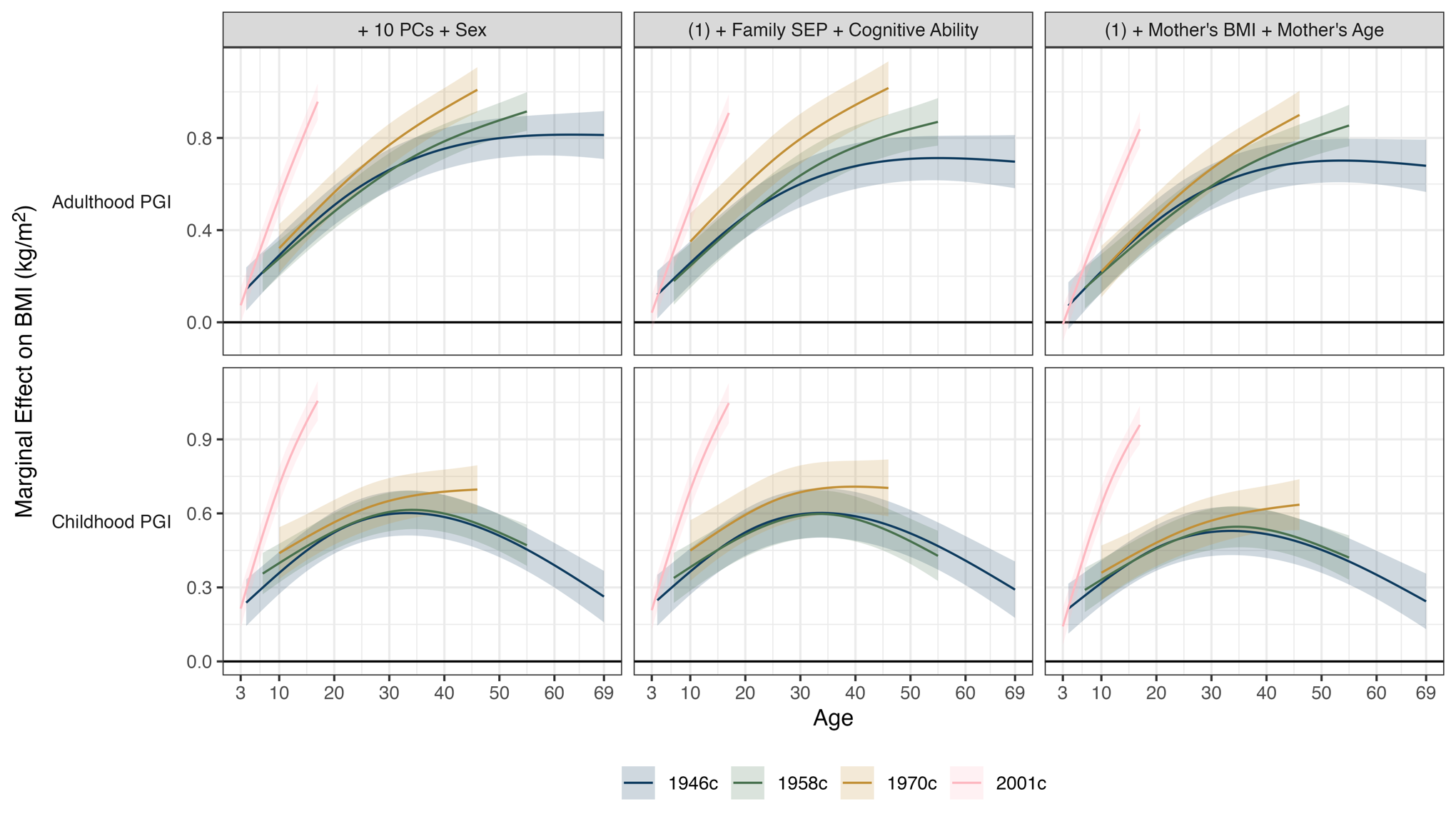

Figure S10: Association between PGI and BMI (kg/m^2^) by cohort, age, PGI (adulthood or childhood) and covariates used. Derived from separate linear mixed effects random intercept models with association between PGI and BMI allowed to vary by age (two natural splines). Left panels: adjustment for sex and first 10 genetic principal components (PCs). Middle panels: adjustment for sex, first 10 genetic PCs, family socioeconomic class, mother’s years of education, and age 10/11 verbal cognitive ability. Right panels: adjustment for sex, first 10 genetic PCs, mother’s BMI, and mother’s age at birth. Estimates were weighted using recruitment weights and account for complex survey design. Estimates were weighted using recruitment weights.

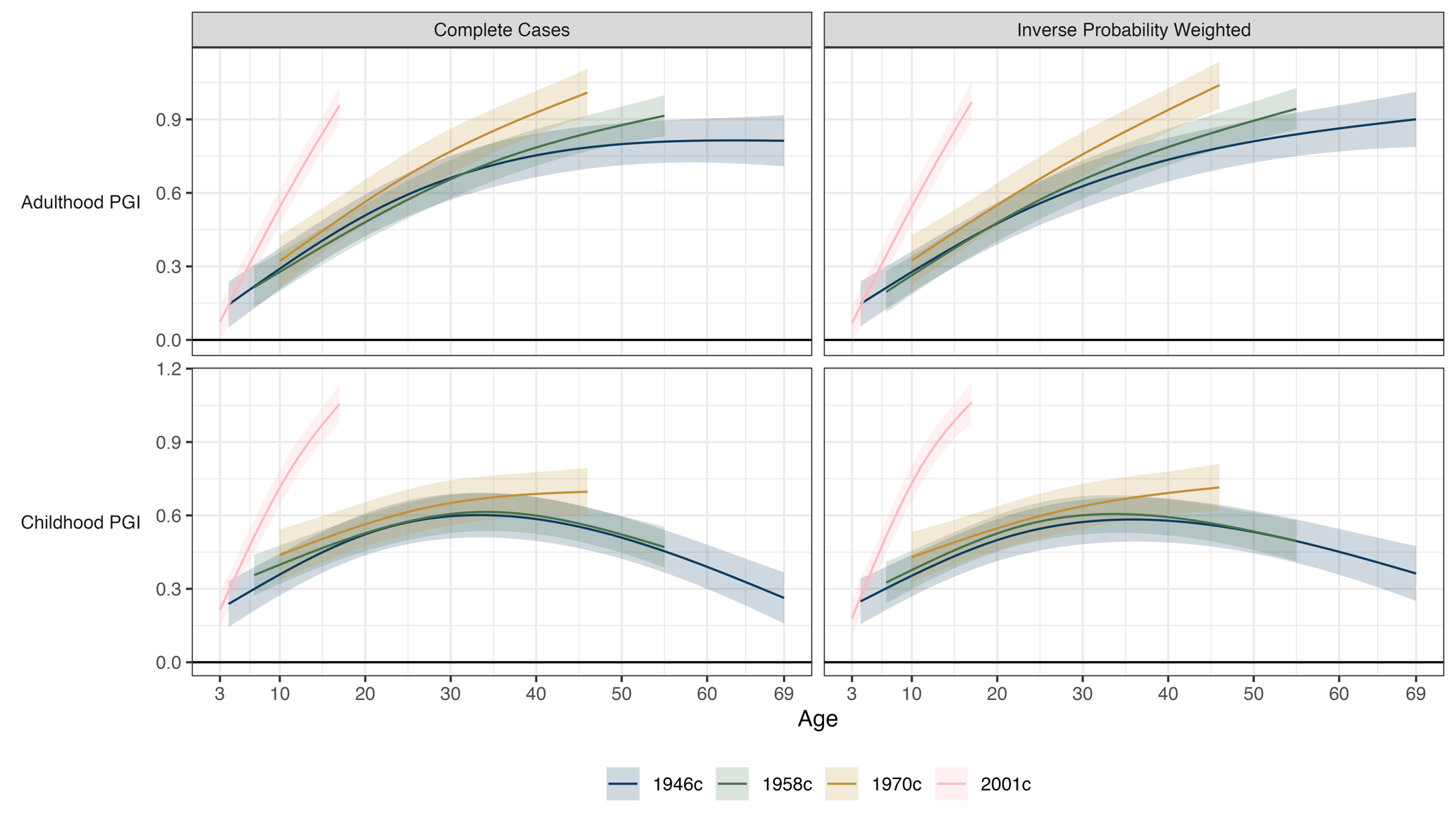

Figure S11: Association between PGI and BMI (kg/m^2^) by cohort, age, PGI (adulthood or childhood) and whether inverse probability weighting (IPW) used to account for selection into genotyped sample. Derived from separate linear mixed effects models with association between PGI and BMI allowed to vary by age (two natural splines). Adjustment for age (two natural splines), sex, first 10 genetic principal components, and person-specific random intercept. Complete case estimates were weighted using recruitment weights. IPW combined these weights with genetic non-response weights.

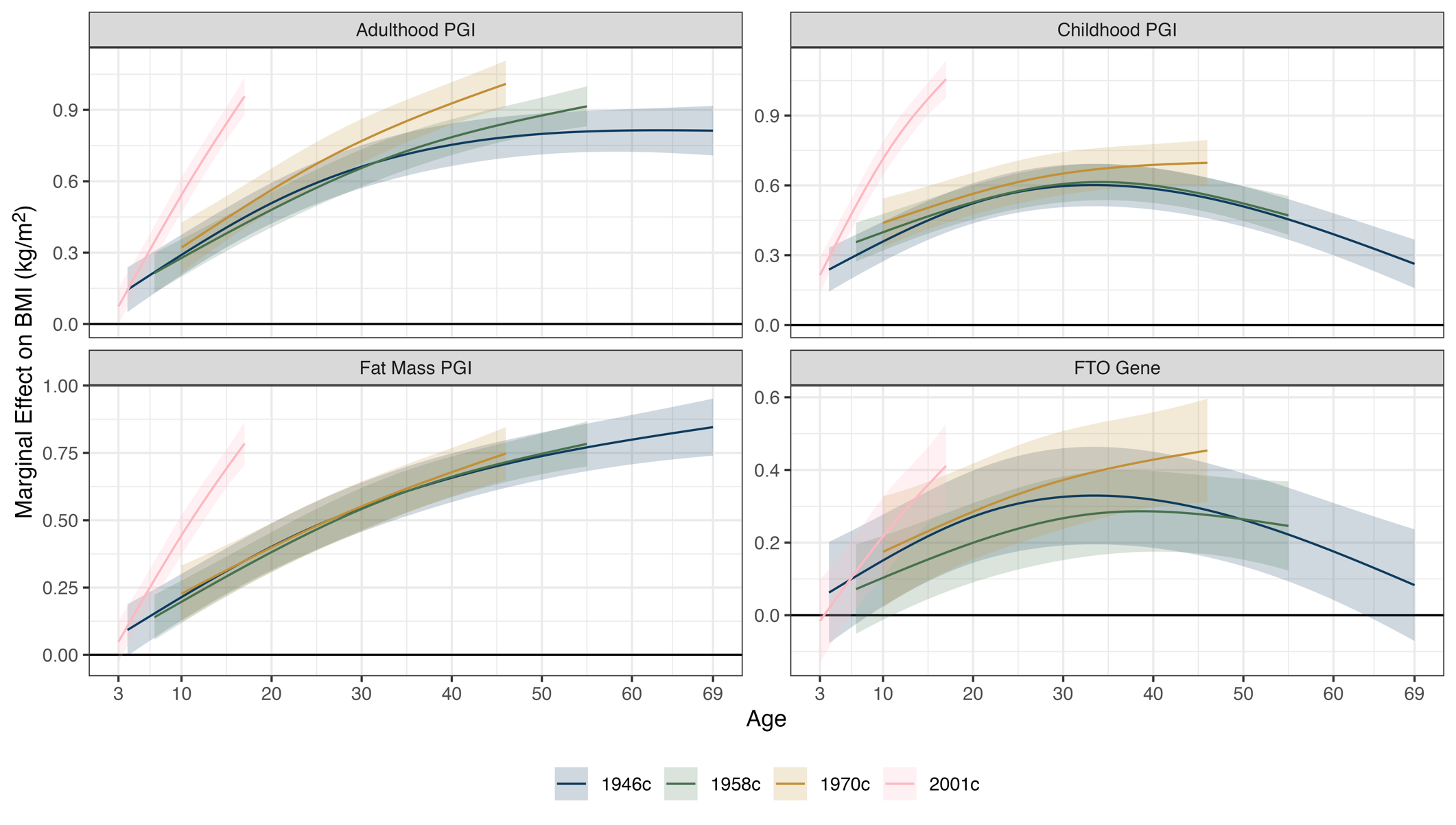

Figure S12: Association between PGI and BMI (kg/m^2^) by cohort, age., and PGI. Derived from separate linear mixed effects models with association between PGI and BMI allowed to vary by age (two natural splines). Adjustment for age (two natural splines), sex, first 10 genetic principal components, and person-specific random intercept. Estimates were weighted using recruitment weights.

### References

Bridges, E. C., Rayner, N. W., Mountford, H. S., Bates, T. C., & Luciano, M. (2023). Longitudinal Reading Measures and Genome Imputation in the National Child Development Study: Prospects for Future Reading Research. *Twin Research and Human Genetics: The Official Journal of the International Society for Twin Studies*, *26*(1), 10–20. https://doi.org/10.1017/thg.2023.2

Centre for Longitudinal Studies. (2024). *CLS Genomics Data*. https://cls-genetics.github.io

Chang, C. C., Chow, C. C., Tellier, L. C., Vattikuti, S., Purcell, S. M., & Lee, J. J. (2015). Second-generation PLINK: Rising to the challenge of larger and richer datasets. *GigaScience*, *4*(1), 7. https://doi.org/10.1186/s13742-015-0047-8

Fitzsimons, E., Moulton, V., Hughes, D. A., Neaves, S., Ho, K., Hemani, G., Timpson, N., Calderwood, L., Gilbert, E., & Ring, S. (2022). Collection of genetic data at scale for a nationally representative population: The UK Millennium Cohort Study. *Longitudinal and Life Course Studies*, *13*(1), 169–187. https://doi.org/10.1332/175795921X16223668101602

Hardy, R., Wills, A. K., Wong, A., Elks, C. E., Wareham, N. J., Loos, R. J. F., Kuh, D., & Ong, K. K. (2010). Life course variations in the associations between FTO and MC4R gene variants and body size. *Human Molecular Genetics*, *19*(3), 545–552. https://doi.org/10.1093/hmg/ddp504

Rubin, D. B. (1987). *Multiple Imputation for Nonresponse in Surveys*. John Wiley & Sons, Ltd. https://doi.org/10.1002/9780470316696

Sullivan, A., Brown, M., Hamer, M., & Ploubidis, G. B. (2023). Cohort Profile Update: The 1970 British Cohort Study (BCS70). *International Journal of Epidemiology*, *52*(3), e179–e186. https://doi.org/10.1093/ije/dyac148
